# A systematic review of the effectiveness of interventions to promote employment rates among people with disabilities in the UK between 2009 and 2024

**DOI:** 10.1101/2024.12.03.24318405

**Authors:** Andy Pennington, David Newbold, Maximilian Limburg, Mary Cronin, Yang Yu, Dorcas N. Magai, Fatemeh Hoda Fallah, Thomas Hampton, Sue Jarvis, John White, Ben Barr, Philip McHale

## Abstract

**Background:** The UK disability employment gap remains at 30 percentage points, with health-related economic inactivity clustered in socioeconomic disadvantaged groups living in places with weak labour markets, costing the economy an estimated £212 billion annually. There is a lack of synthesised UK based evidence of what works for these population groups.

**Objectives:** This systematic review evaluated UK micro- and meso-level employment interventions for people with disabilities to identify effective solutions.

**Methods:** We synthesised quantitative experimental and quasi-experimental UK-based evaluations published between 2009 and 2024 from academic and grey literature.

**Results:** From 8,088 records, 13 studies (14 articles) were included. Individual trial evidence indicates that high-fidelity Individual Placement and Support (IPS) in clinical settings may double competitive employment for severe mental illness (representing 22% vs. 11% in one key trial), and adding motivational interviewing was associated with a more than fourfold increase in employment odds in a localized cluster-RCT (OR=4.3; 95% CI: 1.5, 16.6). For common mental health conditions, co-locating Employment Advisers in psychological therapy (IAPT) was associated with a higher employment rate of 48% (vs. 29% for therapy alone) among clients looking for work in a major evaluation. Mandatory Jobcentre programmes showed no significant employment gains, whereas voluntary schemes like Want2Work were associated with a 7 to 8 percentage point increase in employment probability. Evaluated interventions largely targeted individual employability rather than the work environment, and study participants tended to be better educated and more labour-market-attached than the UK average for people economically inactive due to disability - pointing to major evidence gaps on employer-side adjustments and interventions for more disadvantaged groups.

**Conclusions:** While voluntary, clinically integrated pathways show promise, drawing firm conclusions about ‘what works’ requires caution; these localised findings may need replication across diverse local labour markets. Narrowing the gap may be achieved by shifting from mandatory activation to carefully evaluated, voluntary, employer-focused workplace modifications.

**Research Highlights:** Each of these bullet points is strictly **under the 85-character Elsevier limit** (inclusive of spaces and punctuation):

- UK disability employment gap is 30 percentage points; economic inactivity (including for health reasons) costs £212bn a year.
- Voluntary, clinically integrated pathways significantly increase employment.
- Co-locating advisers in psychological therapy increases job entry rates to 48%.
- Large-scale mandatory Jobcentre activation shows no significant work gains.
- There is a lack of evidence for employer side modifications.

## Background

### The problem and policy context

The rising prevalence of health-related economic inactivity represents a severe social, economic and public health challenge, costing the UK economy an estimated £212 billion annually, or roughly 7% of GDP (Mayfield, 2025). The disability employment gap was 30 percentage points in April–June 2025 (Department for Work and Pensions, 2025c). Around 4 million working-age people in England and Wales now claim health-related benefits, representing a substantial increase since 2019 (Latimer *et al*., 2025). This problem is increasingly driven by mental health conditions, with the share of the working-age population claiming Personal Independence Payment for anxiety and depression nearly tripling between 2012 and 2024, and nearly doubling between February 2020 and August 2024 (Department for Work and Pensions, 2025b). Current UK government policy is attempting to address this challenge through a more support-first-focused approach designed to de-risk the transition into employment, so that people can “try work without fear of losing benefit” (Department for Work and Pensions et al., 2024; Department for Work and Pensions, 2025a). This protects claimants from losing their financial safety net upon taking a job, ensuring they are not left without an income if health or other issues prevent them from sustaining employment. Legislative reforms like the Right to Try Guarantee provide statutory protections so claimants can trial work without the fear of benefit reassessment (Department for Work and Pensions, 2025a, 2026b).

Localised initiatives like the Connect to Work programme and the NHS-partnered WorkWell pilots decentralise support and integrate clinical and vocational care (Department for Work and Pensions et al., 2024; Department for Work and Pensions, 2026a). Standard interventions, however, often fail to account for how poor health and multiple morbidities are heavily clustered among individuals with low skills and low educational attainment in disadvantaged areas with weak local labour markets. Place-based, UK-specific evidence is therefore essential to understand what works for these specific population groups in these places.

The employment of disabled people is closely interrelated with health inequalities. Pronounced health inequalities across the UK mean that poor health and multiple physical and mental health morbidities tend to be highly clustered among people with low skills and education in disadvantaged areas with weak labour markets (Marmot et al., 2010). This clustering contributes to high rates of health-related economic inactivity and disability (Mayfield, 2025). It also means that people who are out of work due to poor health tend to have lower levels of education, for example in 2019, only 8% of people economically inactive due to long-term sickness had a degree-level qualification (Office for National Statistics, 2019), a high prevalence of common mental health problems - often co-occurring with physical illness - and tend to live in areas where employment opportunities are scarcer and frequently characterised by poor quality, insecure work (Bambra et al., 2009). Being out of work in turn increases the risk of poverty, which can further worsen health and deepen these inequalities, creating a self-reinforcing cycle (Waddell and Burton, 2006; Marmot et al., 2010). Evidence of what works needs to take account of the complex place-based context of health-related economic inactivity in the UK. Despite decades of policies aiming to promote the employment of people with disabilities, and multiple reviews of the evidence, substantial gaps remain – we do not know what works – for these population groups in these places (Clayton et al., 2011, 2012; Vooijs et al., 2015).

Interventions aiming to reduce the disability employment gap can work at various levels. Micro-level interventions are individual-focused strategies designed to strengthen personal capabilities and enhance the direct "employability" of disabled or chronically ill individuals, such as through personal adviser support, vocational training, and targeted financial incentives. Conversely, meso-level interventions are organisational- or employer-focused, aiming to adapt the immediate working environment, modify job design, or alter employer behaviours, including implementing physical workspace adjustments and offering flexible schedules. Lastly, macro-level interventions operate as broad, societal-wide structural levers that reshape national legislative, welfare, or economic frameworks to affect the entire population, such as through statutory anti-discrimination acts or national benefit structures (Clayton *et al*., 2011; Whitehead *et al*., 2009).

### What is already known and why another review is needed

Several previous systematic and umbrella reviews have assessed the effectiveness of disability employment interventions, but each has clear limitations. Bambra et al. (2009) showed that organisational-level changes to the psychosocial work environment can improve employee health and reduce inequalities, but the underlying studies lacked causally robust designs. Clayton et al. (2011) evaluated individual-focused UK return-to-work initiatives and found that personal adviser models helped some claimants, but were undermined by high adviser workloads and output targets that incentivised "cream-skimming" of work-ready claimants while neglecting those furthest from the labour market. Clayton et al. (2012) reviewed employer-focused initiatives and concluded that workplace adjustments had positive impacts on employment but low uptake, that some interventions favoured more advantaged disabled people and those closer to the labour market, and that participant selection may bias even controlled studies. Vooijs et al. (2015) evaluated generic work-directed interventions across chronic diseases and found that interventions directed at the work environment were mostly reported as effective.

Several systematic reviews of randomised controlled trials (RCTs) have shown that the Individual Placement and Support (IPS) model is effective at increasing employment among people with severe mental illness (Modini et al., 2016; Suijkerbuijk et al., 2017), including young adults with serious mental illness or early psychosis (Bond et al., 2023). Whitworth et al. (2024) found IPS more effective than business-as-usual at supporting disabled people without severe mental illness into employment, but only two of the included studies were from the UK (Marsden et al., 2024; Newton et al., 2023). Bond et al. (2026) found no significant effect for IPS with common mental disorders in a meta-analysis of five studies (none from the UK), but did find improved employment from IPS for people with substance misuse disorders, from a meta-analysis of five studies (one from the UK).

Limiting evidence reviews to RCTs may be too restrictive, as this tends to exclude large national welfare-to-work initiatives aimed at broad client groups; however, including uncontrolled evaluations introduces bias from the selection of work-ready participants into programmes, as outlined above. Previous reviews have therefore fallen short by failing to include a broad range of causal study designs while systematically assessing the extent to which studies include more disadvantaged groups, such as those with low skills or long-term worklessness. A new systematic review is therefore needed to synthesise higher-quality UK evidence using robust causal counterfactuals, and to evaluate how well the existing evidence addresses the socioeconomic disadvantage associated with health-related economic inactivity in the UK.

We, therefore, aimed to systematically review the evidence from experimental and quasi-experimental studies for effectiveness of micro and meso level interventions designed to increase the employment of people with disabilities in the UK comparing the labour market attachment and educational level of intervention participants to the average for people out of work with disabilities in the UK.

The aim of this systematic review was to answer two questions:

**RQ1** What is the higher methodological quality evidence (from experimental and quasi-experimental, studies with control groups) on the effectiveness of micro- and meso-level interventions designed to increase employment rates among people with disability in the UK between 2009 and 2024?
**RQ2** To what extent was this interventional evidence derived from, and representative of people out of work due to disability in the UK?

## Methods

Approaches in the review were informed by the Centre for Reviews and Dissemination’s guidance (Centre for Reviews and Dissemination, 2009). A systematic review protocol was registered with Prospero (registration number: CRD42018103930). Reporting complies with the Preferred Reporting Items for Systematic Reviews and Meta-Analyses (PRISMA) 2020 statement (Page et al., 2021) and its Equity Extension (PRISMA-Equity) guidelines (Welch et al., 2012); the completed compliance checklists are provided in Appendix 7.

### Study identification

Inclusion and exclusion criteria for the review are shown in Table 1.

**Table 1.** Systematic review inclusion and exclusion criteria.

|  | Include | Exclude |
| --- | --- | --- |
| Population and setting | Individuals with disability (including limiting long-term conditions); UK. | Studies that aggregate/conflate results for individuals with a disability with other population groups. |
| Studies of interest | Quantitative longitudinal or repeated cross section studies with control groups. | Studies without an appropriate comparator/control group. |
| Intervention type | Micro- (individual) and meso- (organisational) level interventions | Macro- (societal, economic, legislative) level interventions (e.g., |
|  | to increase levels of employment rates. | legislation/Acts of Parliament banning workplace discrimination). |
| Outcomes | Outcomes measuring levels of employment levels/rates. | - |
| Publication Types | Complete peer reviewed academic empirical articles, grey literature reports. | Pre-print articles, non-empirical, e.g., opinion or discussion pieces, conference abstracts or posters.<br>Incomplete publications, not containing reporting of methods and results. |
| Time coverage | Published between 2009 and 2024. | - |
| Language | English-language studies. | - |

This review was on evidence from quantitative studies of effectiveness. Only studies with control/comparison groups were included to ensure there was the necessary counterfactual to determine the level of effect of the intervention. The review also focussed specifically on UK to identify evidence from studies in similar geographical, sociocultural, and policy contexts to the UK. The review sought to locate and synthesise evidence on micro- and meso-level interventions that are aimed at: i. ‘strengthening’ individuals/improving the ‘employability’ of people with disabilities themselves, and ii. improving the employment environment for individuals (Whitehead et al., 2009).

### Searches

To systematically identify quantitative evaluations of micro- and meso-level disability employment interventions in the UK published between 2009 and 2024, we deployed a dual-stream search strategy targeting both peer-reviewed academic literature and grey literature. Given that UK government departments frequently commission welfare-to-work evaluations that are published exclusively in grey formats, we implemented a rigorous grey literature search protocol first. This consisted of advanced, date-limited Google and web domain-targeted (.gov.uk and organisational) searches using Boolean search operators, hand searches of specialist repositories, and targeted artificial intelligence (AI) search engines (ChatGPT 4o, MS Copilot) to ensure saturation. Once saturation was achieved, we translated the search strategy into specific syntaxes and executed them across five ‘gold-standard’ academic bibliographic databases: OVID MEDLINE (including Epub Ahead of Print and In-Process Citations), Web of Science Social Sciences Citation Index (SSCI), APA PsycInfo, CINAHL Ultimate, and EconLit. Literature screening was conducted independently by two reviewers at both the title/abstract and full-text stages. To maximise completeness, we supplemented these streams with forward and backward citation snowballing of all included publications utilising Web of Science, Research Rabbit AI, and Google Scholar, alongside direct contact with topic experts. Non-English and pre-2009 studies, as well as those lacking a robust quantitative or statistical counterfactual control group, were excluded.

The use of AI in any/all stages of this review were informed by the Position statement on artificial intelligence (AI) use in evidence synthesis across Cochrane, the Campbell Collaboration, JBI and the Collaboration for Environmental Evidence 2025 (Flemyng *et al*., 2025). In the absence of PRISMA guidance on reporting of the use of AI in systematic review, to ensure transparency and replicability, a detailed description of the approaches is in Appendix 1, together with an example of the (MEDLINE) search strategy.

### Screening

Academic publications were selected for inclusion in two stages: title and abstract screening, and full text screening. Titles and abstracts were screened independently by two reviewers. Articles and reports deemed potentially eligible for inclusion were then retrieved and screened independently by two reviewers as full texts. Screening of academic publications was conducted in EPPI-Reviewer 6 systematic review management software which uses compatible export files containing results from databases (Thomas *et al*., 2023). This was not possible for grey literature search results as Google does not provide such files. Google search results were therefore screened manually and independently by two reviewers who followed each initial result deemed relevant through to full text sources at each stage of the searches described above.

Screening of each Google search was limited to the first 100 results because the relevance of the search results diminishes rapidly due to prioritisation algorithms (Google, 2025). Lists of included studies, and reasons for exclusion of full texts were recorded in accordance with PRISMA guidelines (Page *et al*., 2021) (Appendix 4). Queries and disagreements on inclusion/exclusion were resolved by consensus or by recourse to a third reviewer.

### Data extraction

Data from each included academic publication was extracted into pre-designed and piloted tables. To address the logistical challenge of extracting data from the larger grey literature publications, data identification and extraction from grey literature publications was aided by use of AI software (Gemini Notebook, Claude). Data extraction from extensive grey literature reports was systematically aided by AI software (Google Gemini Notebook) under strict human reviewer verification. Relevant data was located in the documents using a series of manually administered prompts/questions, and when required additional prompts, by an experienced systematic reviewer (Appendix 1). AI responses were checked individually by the reviewer who maintained human oversight throughout. All data extraction tables were completed by one reviewer and then checked for accuracy and completeness independently by a second reviewer. Queries and disagreements were resolved by discussion, or by recourse to a third reviewer. To compare the study populations to the UK population out of work due to disability (including chronic illness) we extracted data form studies on participants educational qualifications and employment history. For comparison we analysed data from the 2019 Annual Population Survey (Office for National Statistics, 2019) for all people economically inactive for long-term sickness. This is the largest economically inactive group in the UK accounting for 5% of the working age population. For interventions to be effective at reducing the disability employment gap, they would need to be effective for this group.

### Methodological quality assessment

Methodological quality of included studies was assessed using a modified version of a tool designed and tested for effectiveness in systematic reviews (Orton *et al*., 2016; Appendix 2). Methodological quality assessments (QAs) were conducted independently by two reviewers. Queries and disagreements were resolved by discussion, or by recourse to a third reviewer.

### Data synthesis

Results were narratively synthesised. In line with guidance and based on the findings of the methodological quality assessment, higher methodological studies are reported first and in greater detail where appropriate (Popay et al., 2006). The potential for meta-analysis of the included study’s findings was explored. It was not appropriate due to high levels of heterogeneity (study intervention types, outcome measures, methods of analysis).

The primary effect measures extracted and evaluated across the quantitative studies were Odds Ratios (OR), Risk/Relative Risks (RR) or percentage point differences.

Due to the high clinical and methodological heterogeneity across the included evaluations, statistical assessments of reporting bias (such as funnel plots) were not feasible. Instead, reporting and publication bias were minimised methodologically through a rigorous, phased grey literature search strategy targeting UK government domains (.gov.uk) and specialist organisational repositories. This protocol was explicitly designed to capture extensive, government-commissioned evaluations that are frequently withheld from academic journal publication and are thus absent from bibliographic databases. Saturation was verified by cross-referencing DWP-commissioned pilot identifiers using conversational AI engines, ensuring that evaluations with null or mixed findings were fully integrated to counter academic publication bias.

## Results

### Results of the literature search

From 8,088 unique records, 14 articles (from 13 studies) studies meeting our inclusion/exclusion criteria were included (Figure 1).

**Figure 1.**
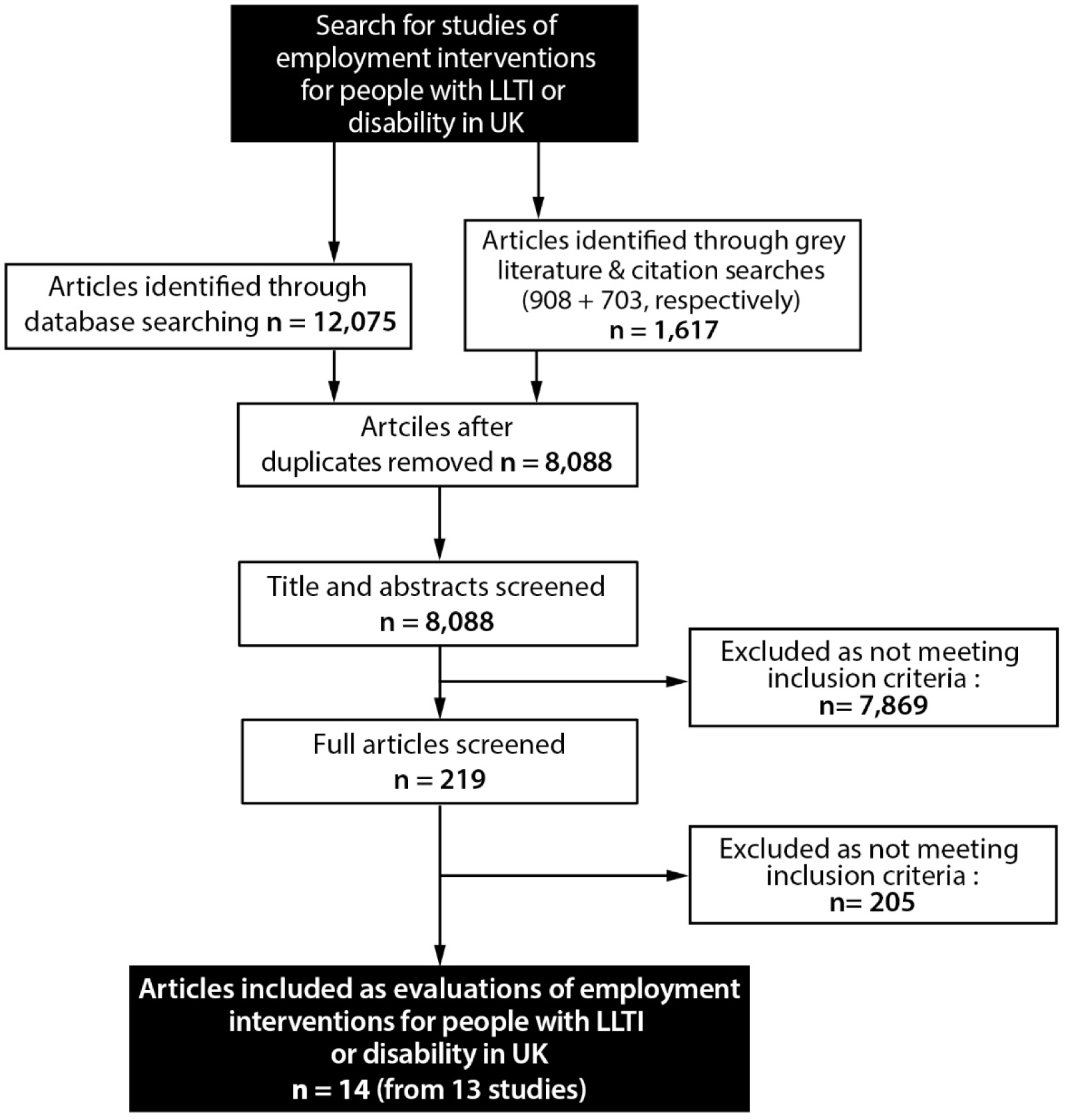
PRISMA flow chart of the progression of studies through the review.

Reasons for excluding studies during full text screening are shown in Appendix 4. A list of the included articles is contained within Appendix 5.

### Characteristics of included studies

Key characteristics of the studies are shown in Table 2 and summarised below.

**Table 2.**
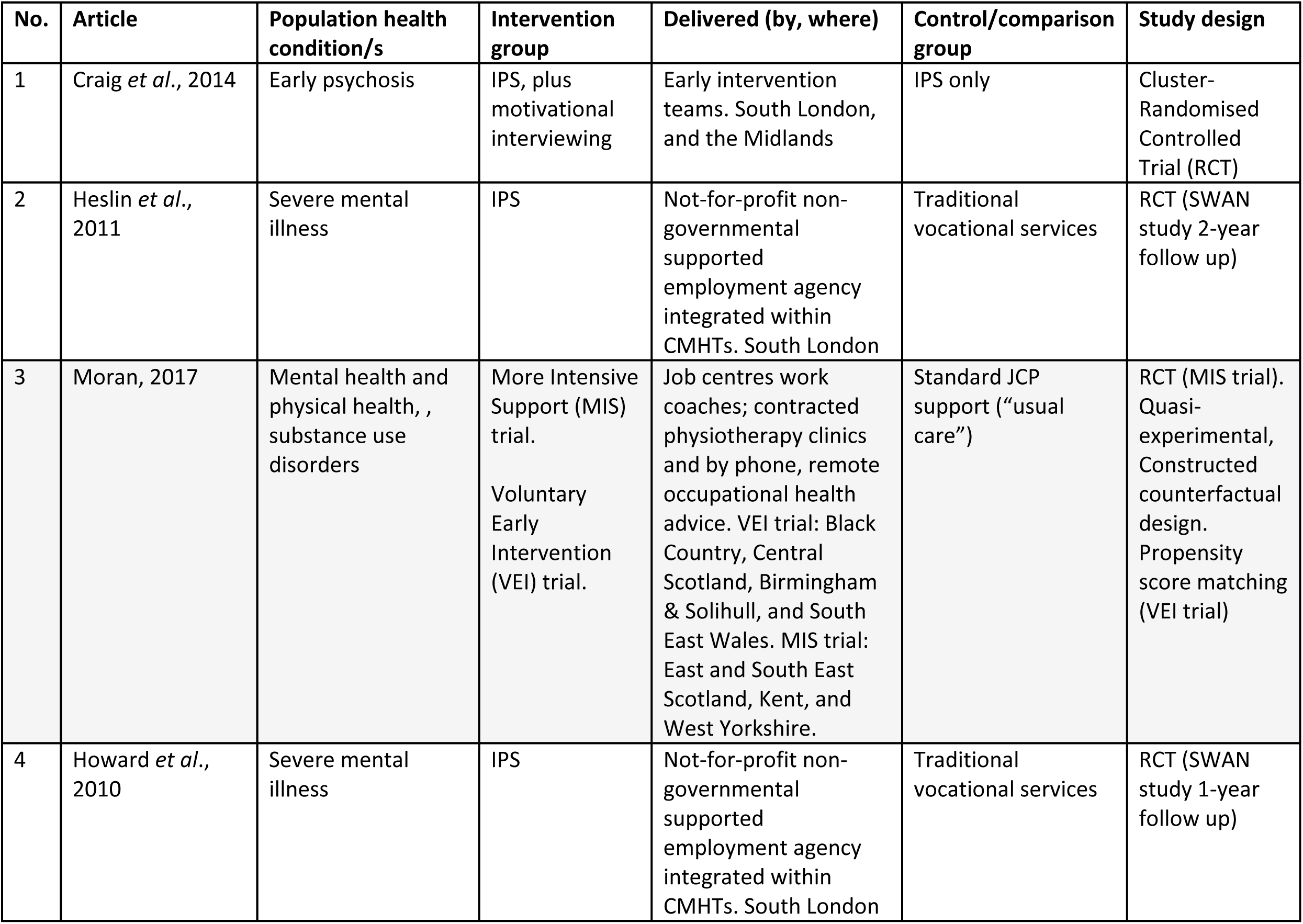
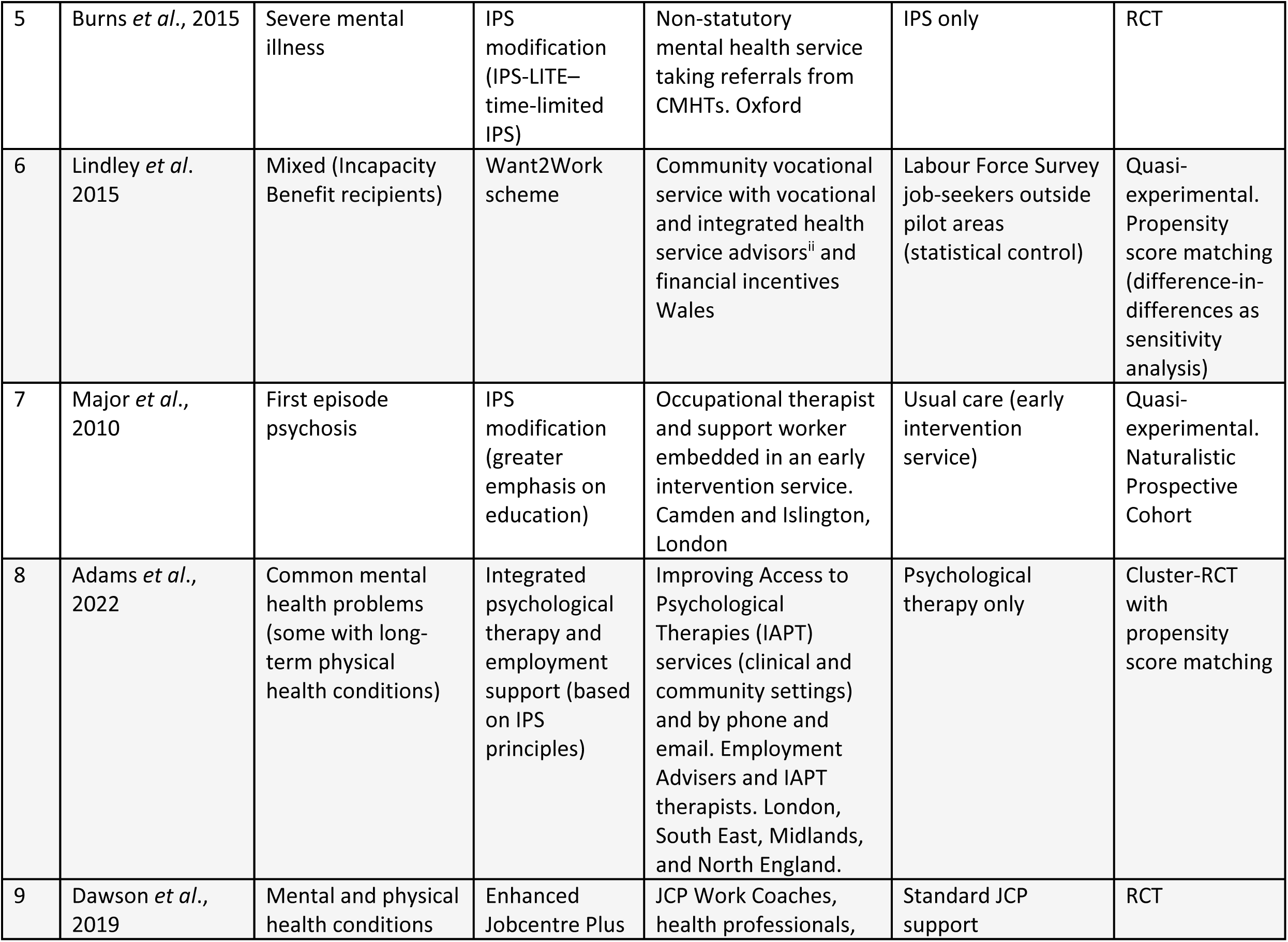
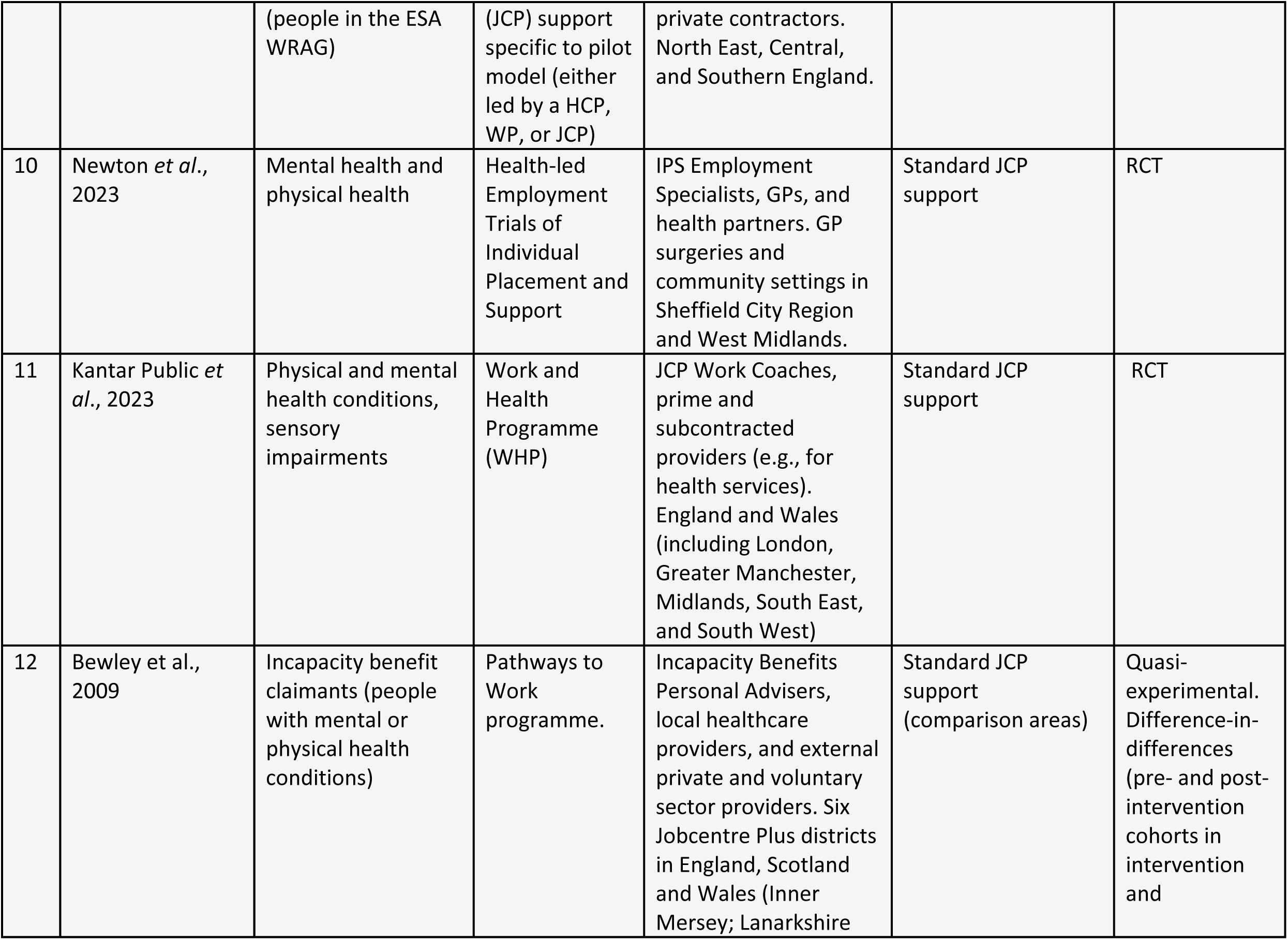
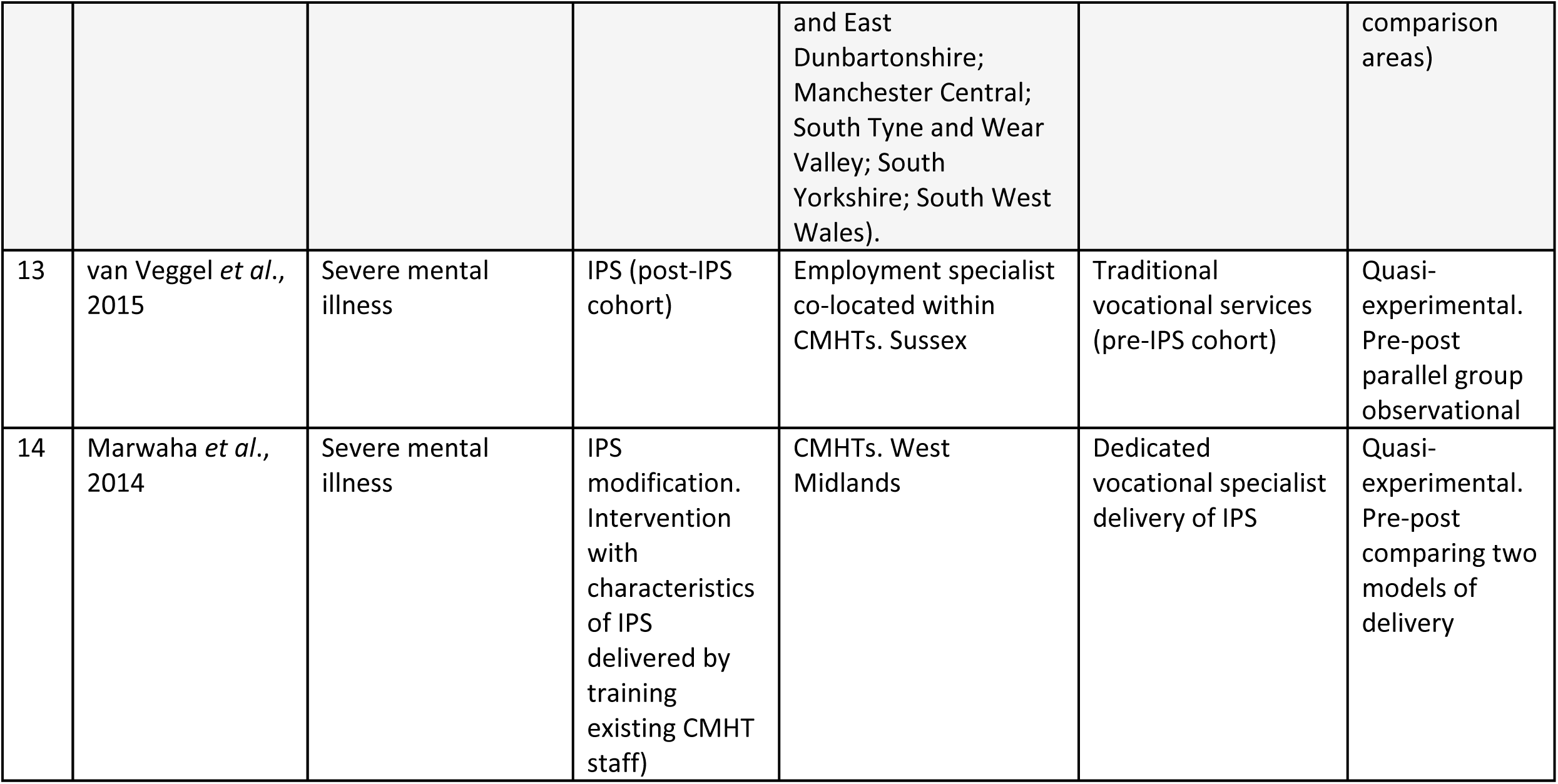
Characteristics of included studies.

| **No.** | **Article** | **Population health condition/s** | **Intervention group** | **Delivered (by, where)** | **Control/comparison group** | **Study design** |
| --- | --- | --- | --- | --- | --- | --- |
| 1 | Craig *et al.*, 2014 | Early psychosis | IPS, plus motivational interviewing | Early intervention teams. South London, and the Midlands | IPS only | Cluster-Randomised Controlled Trial (RCT) |
| 2 | Heslin *et al.*, 2011 | Severe mental illness | IPS | Not-for-profit non-governmental supported employment agency integrated within CMHTs. South London | Traditional vocational services | RCT (SWAN study 2-year follow up) |
| 3 | Moran, 2017 | Mental health and physical health, , substance use disorders | More Intensive Support (MIS) trial.<br>Voluntary Early Intervention (VEI) trial. | Job centres work coaches; contracted physiotherapy clinics and by phone, remote occupational health advice. VEI trial: Black Country, Central Scotland, Birmingham & Solihull, and South East Wales. MIS trial: East and South East Scotland, Kent, and West Yorkshire. | Standard JCP support ("usual care") | RCT (MIS trial). Quasi-experimental, Constructed counterfactual design. Propensity score matching (VEI trial) |
| 4 | Howard *et al.*, 2010 | Severe mental illness | IPS | Not-for-profit non-governmental supported employment agency integrated within CMHTs. South London | Traditional vocational services | RCT (SWAN study 1-year follow up) |
| 5 | Burns *et al.*, 2015 | Severe mental illness | IPS modification (IPS-LITE–time-limited IPS) | Non-statutory mental health service taking referrals from CMHTs. Oxford | IPS only | RCT |
| 6 | Lindley *et al.* 2015 | Mixed (Incapacity Benefit recipients) | Want2Work scheme | Community vocational service with vocational and integrated health service advisors<sup>ii</sup> and financial incentives Wales | Labour Force Survey job-seekers outside pilot areas (statistical control) | Quasi-experimental. Propensity score matching (difference-in-differences as sensitivity analysis) |
| 7 | Major *et al.*, 2010 | First episode psychosis | IPS modification (greater emphasis on education) | Occupational therapist and support worker embedded in an early intervention service. Camden and Islington, London | Usual care (early intervention service) | Quasi-experimental. Naturalistic Prospective Cohort |
| 8 | Adams *et al.*, 2022 | Common mental health problems (some with long-term physical health conditions) | Integrated psychological therapy and employment support (based on IPS principles) | Improving Access to Psychological Therapies (IAPT) services (clinical and community settings) and by phone and email. Employment Advisers and IAPT therapists. London, South East, Midlands, and North England. | Psychological therapy only | Cluster-RCT with propensity score matching |
| 9 | Dawson *et al.*, 2019 | Mental and physical health conditions | Enhanced Jobcentre Plus | JCP Work Coaches, health professionals, | Standard JCP support | RCT |
|  |  | (people in the ESA WRAG) | (JCP) support specific to pilot model (either led by a HCP, WP, or JCP) | private contractors. North East, Central, and Southern England. |  |  |
| 10 | Newton *et al.*, 2023 | Mental health and physical health | Health-led Employment Trials of Individual Placement and Support | IPS Employment Specialists, GPs, and health partners. GP surgeries and community settings in Sheffield City Region and West Midlands. | Standard JCP support | RCT |
| 11 | Kantar Public *et al.*, 2023 | Physical and mental health conditions, sensory impairments | Work and Health Programme (WHP) | JCP Work Coaches, prime and subcontracted providers (e.g., for health services). England and Wales (including London, Greater Manchester, Midlands, South East, and South West) | Standard JCP support | RCT |
| 12 | Bewley *et al.*, 2009 | Incapacity benefit claimants (people with mental or physical health conditions) | Pathways to Work programme. | Incapacity Benefits Personal Advisers, local healthcare providers, and external private and voluntary sector providers. Six Jobcentre Plus districts in England, Scotland and Wales (Inner Mersey; Lanarkshire | Standard JCP support (comparison areas) | Quasi-experimental. Difference-in-differences (pre- and post-intervention cohorts in intervention and |
|  |  |  |  | and East Dunbartonshire; Manchester Central; South Tyne and Wear Valley; South Yorkshire; South West Wales). |  | comparison areas) |
| 13 | van Veggel *et al.*, 2015 | Severe mental illness | IPS (post-IPS cohort) | Employment specialist co-located within CMHTs. Sussex | Traditional vocational services (pre-IPS cohort) | Quasi-experimental. Pre-post parallel group observational |
| 14 | Marwaha *et al.*, 2014 | Severe mental illness | IPS modification. Intervention with characteristics of IPS delivered by training existing CMHT staff) | CMHTs. West Midlands | Dedicated vocational specialist delivery of IPS | Quasi-experimental. Pre-post comparing two models of delivery |

There were two main groups of studies: First, relatively small trials of individual placement and support (IPS) interventions published in peer-reviewed academic journals; these were mainly RCTs (Burns *et al*., 2015; Craig *et al*., 2014; Heslin *et al*., 2011; Howard *et al*., 2010; Major *et al*., 2010; Marwaha *et al*., 2014; van Veggel *et al*., 2015). These studies are highlighted as grey rows in Tables 2 and 4. Second, evaluations of large existing government employment programmes or pilots (trials) commission by government departments and published as grey literature (Adams *et al*., 2022; Bewley *et al*., 2009; Dawson *et al*., 2019; Kantar Public *et al*., 2023; Moran, 2017; Newton *et al*., 2023); with Lindley *et al*. (2015) as the only exception being published in a peer-reviewed academic journal. These studies are shown in the white rows of Tables 2 and 4.

Most of the IPS trials either investigated standard (high fidelity) models of IPS or modified versions thereof, delivered through integration of employment support within Community Mental Health Teams (CMHTs), with one exception that trained CMHT staff to deliver IPS (Marwaha *et al*., 2014). All but two of the trials of IPS interventions included participants with severe mental health conditions. Exceptions were Newton *et al*.’s (2023) evaluation of the Health-led Employment Trials of IPS with participants with a range of physical and self-defined low to moderate mental health conditions (a shift from traditional IPS), and Adams *et al*.’s (2022) evaluation of the Employment Advisers in Improving Access to Psychological Therapies intervention that was based on IPS principles. The evaluations of government programmes included participants with a range of mental and physical conditions and disabilities, and sometimes substance use disorders. They were delivered across a variety of settings shown in Table 2. Across all the studies employment outcomes were measured through employment rates in either competitive or voluntary employment (Table 4). Eight utilised Randomised Control Trial (RCT) or cluster-RCT study designs, five used various quasi-experimental designs, and one (Moran, 2017) incorporated both RCT and quasi-experimental designs.

### Overview of the employment interventions in the included studies

#### Individual placement and support (IPS)

IPS was originally developed to support people with severe mental illness (e.g., schizophrenia, bipolar disorder) into employment (Bond, 2004; Drake, Bond and Becker, 2012). It follows a "place first, train later" approach, in contrast to traditional "train first, place later" vocational models (Bond, Drake and Becker, 2008), and rests on eight principles: (1) focus on open/competitive employment; (2) open to all who want to work, irrespective of condition; (3) rapid job search; (4) jobs matched to individual preference; (5) employment specialists integrated into clinical teams; (6) employer relationships built around the person’s preferences; (7) ongoing individualised support for both person and employer; and (8) integrated benefits counselling (Becker and Drake, 2003; Bond, 2004; Drake, Bond and Becker, 2012). Interventions that closely follow these principles are termed "high fidelity" IPS.

#### Government employment programmes

The included studies evaluated seven government employment programmes or pilots, summarised below by their distinguishing features.

The **2015 ESA trials** (Moran, 2017) covered three models (only VEI and MIS had impact estimates; the claimant commitment strand was qualitative): *voluntary early intervention* (∼6 hours of personalised work-coach time at the start of an ESA claim, using behavioural-insight techniques); the *claimant commitment* (structured, goal-oriented coaching conversations); and *more intensive support* (mandatory, for Work Programme completers still without a job, offering substantially increased adviser time).

##### Want2Work

(Lindley et al., 2015) was a voluntary scheme for Incapacity Benefit recipients combining community-based advisers, integrated health guidance, financial incentives (e.g., job-preparation premiums), training funding, and in-work support during the first year of employment.

##### Employment Advisers in IAPT

(Adams et al., 2022) embedded employment advisers within mental health services for people with common mental health conditions, offering flexible timing relative to psychological treatment, practical job-search help, and work on participants’ beliefs about their own capability.

##### Pathways to Work

(Bewley et al., 2009) combined mandatory work-focused interviews with a voluntary "choices" package: a healthcare-professional-led condition-management programme, specialised disability employment support, work-preparation courses, and work trials.

The **ESA Work-Related Activity Group pilots** (Dawson et al., 2019) tested three delivery models within a mandatory two-year framework: healthcare-provider-led (mandatory health-focused meetings), Work Programme-led (private-provider discretion, "black box" delivery), and Jobcentre Plus-led (additional Work Coach time and training).

The **Health-led Employment Trials** (Newton et al., 2023) applied an IPS-based model for people with self-defined mild-to-moderate physical or mental health conditions, delivered voluntarily by specialists embedded in primary care: nine months of intensive job search plus four months of in-work support, alongside employer engagement and health referrals.

The **Work and Health Programme** (Kantar Public et al., 2023) supported disabled people and other priority groups (e.g., long-term unemployed) through a dedicated key worker for up to 15 months, offering job-search help, skills training, financial assistance with job-related costs, coordination of specialist health/addiction support, and up to 6 months of in-work follow-up.

### Study methodological quality assessment (QA)

The results of the methodological QA are shown in Table 3.

**Table 3.** Summary of methodological quality assessment (QA) results - scores and ranking.

| No. | Paper (author, year) | Selection bias<br>A to C (3 to 1) | Study design<br>A to D (3 to 0) | Confounders<br>A to C (3 to 1) | Blinding<br>A to C (3 to 1) | Data collection<br>A to C (3 to 1) | Withdrawals & dropouts<br>A to C (3 to 1) | SCORE<br>(Max 18) | QA level |
| --- | --- | --- | --- | --- | --- | --- | --- | --- | --- |
| 1 | Craig *et al.*, 2014 | A (3) | A (3) | A (3) | B (2) | A (3) | A (3) | 17 | Very high |
| 2 | Heslin *et al.*, 2011 | A (3) | A (3) | A (3) | B (2) | A (3) | A (3) | 17 |  |
| 3 | Moran, 2017 | A (3) | A (3) | A (3) | B (2) | A (3) | A (3) | 17 |  |
| 4 | Howard *et al.*, 2010 | B (2) | A (3) | A (3) | B (2) | A (3) | A (3) | 16 |  |
| 5 | Burns *et al.*, 2015 | A (3) | A (3) | A (3) | C (1) | A (3) | B (2) | 15 | High |
| 6 | Lindley *et al.*, 2015 | C (1) | A (3) | A (3) | A (3) | A (3) | B (2) | 15 |  |
| 7 | Major *et al.*, 2010 | C (1) | A (3) | A (3) | C (1) | A (3) | A (3) | 14 |  |
| 8 | Adams *et al.*, 2022 | C (1) | A (3) | A (3) | C (1) | A (3) | B (2) | 13 | Medium |
| 9 | Dawson *et al.*, 2019 | B (2) | A (3) | B (2) | C (1) | A (3) | B (2) | 13 |  |
| 10 | Newton *et al.*, 2023 | B (2) | A (3) | A (3) | C (1) | A (3) | C (1) | 13 |  |
| 11 | Kantar Public *et al.*, 2023 | C (1) | A (3) | A (3) | C (1) | A (3) | C (1) | 12 |  |
| 12 | Bewley *et al.*, 2009 | C (1) | B (2) | A (3) | C (1) | C (1) | A (3) | 11 |  |
| 13 | van Veggel *et al.*, 2015 | C (1) | B (2) | A (3) | C (1) | C (1) | C (1) | 9 | Low |
| 14 | Marwaha *et al.*, 2014 | C (1) | A (3) | C (1) | C (1) | C (1) | C (1) | 8 |  |
|  |  | 28/48 | 46/48 | 44/48 | 24/48 | 42/48 | 35/48 |  |  |

This QA included a critical appraisal and a relative ranking of studies that are of inherently higher quality designs. Although all included studies used control groups, we identified variations in methodological quality attributable to differences in individual study designs, operationalisation, and reporting. Of 14 articles (from 13 studies), half (7) scored as high (3) or very high quality (4), five as medium quality, and two as relatively low quality. Two of the evaluations of larger-scale publicly funded employment programmes or pilots were of higher quality (Moran, 2017, very high; Lindley et al., 2015, high). The other five were all rated as medium quality. Looking across the studies there appears to have been weaknesses in achieving representative selection of participants at baseline, in blinding (of participants, intervention deliverers, or data analysts), and with withdrawals and dropouts during delivery.

### Study findings

Fourteen UK articles from 13 studies evaluated employment outcomes. Findings from the studies are summarised in Table 4 below, ordered by methodological quality (higher to lower).

**Table 4.**
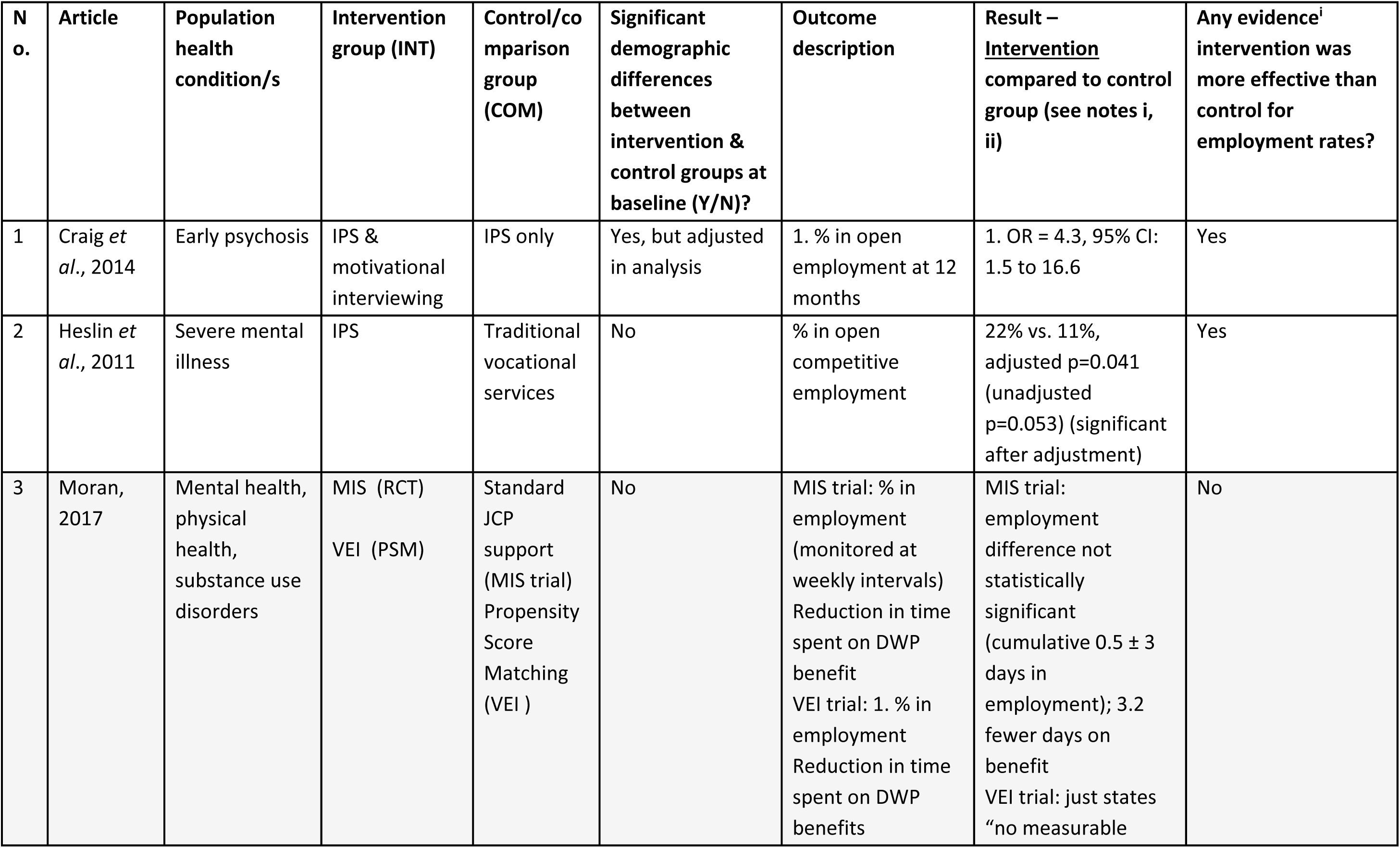
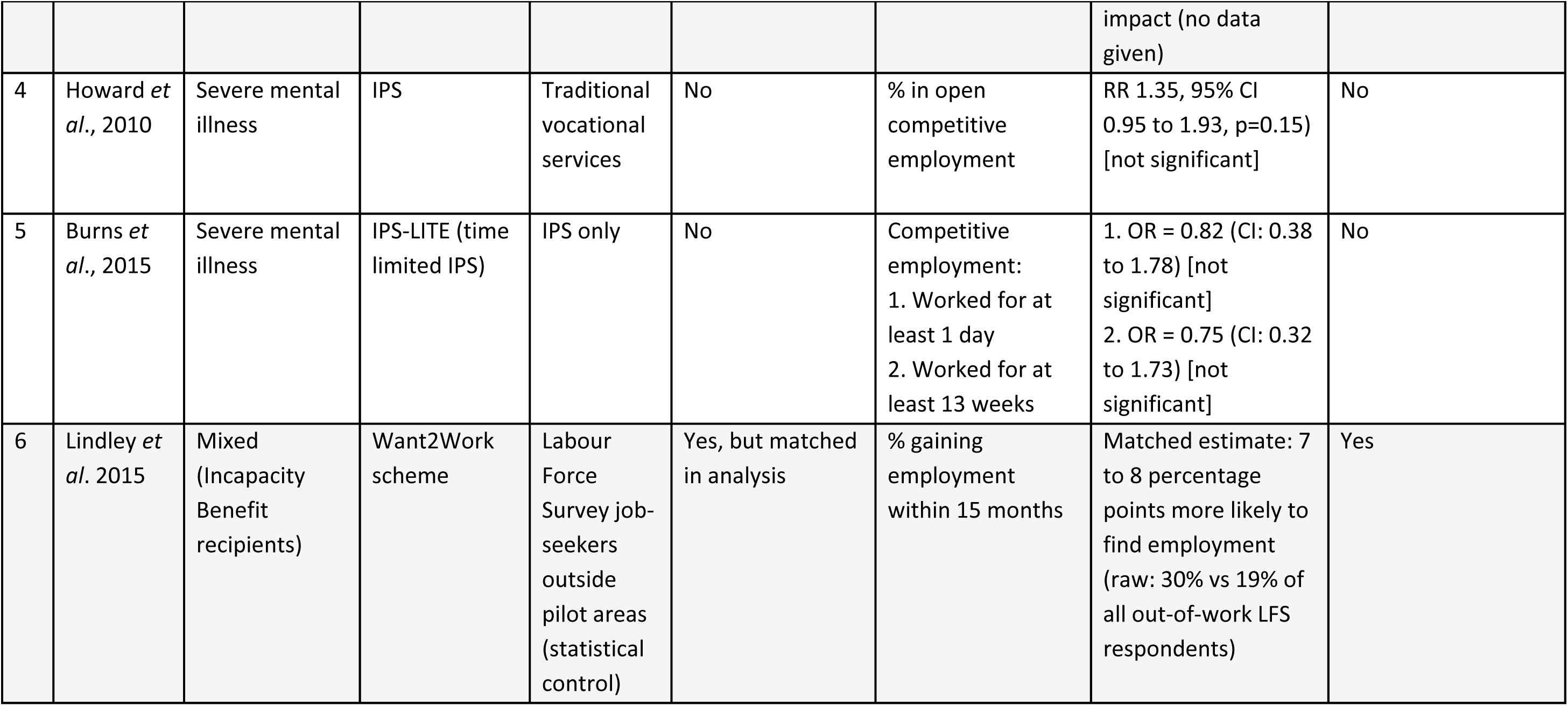
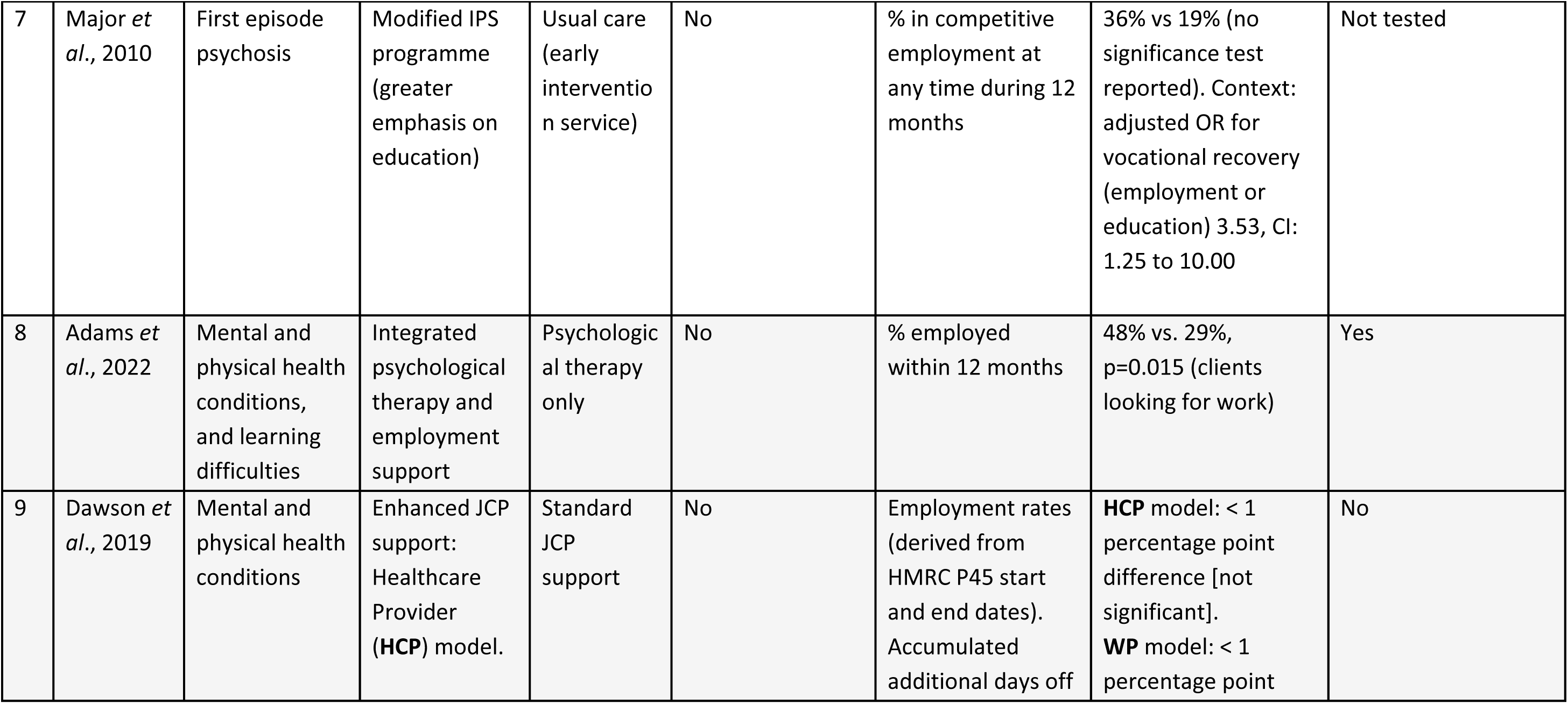
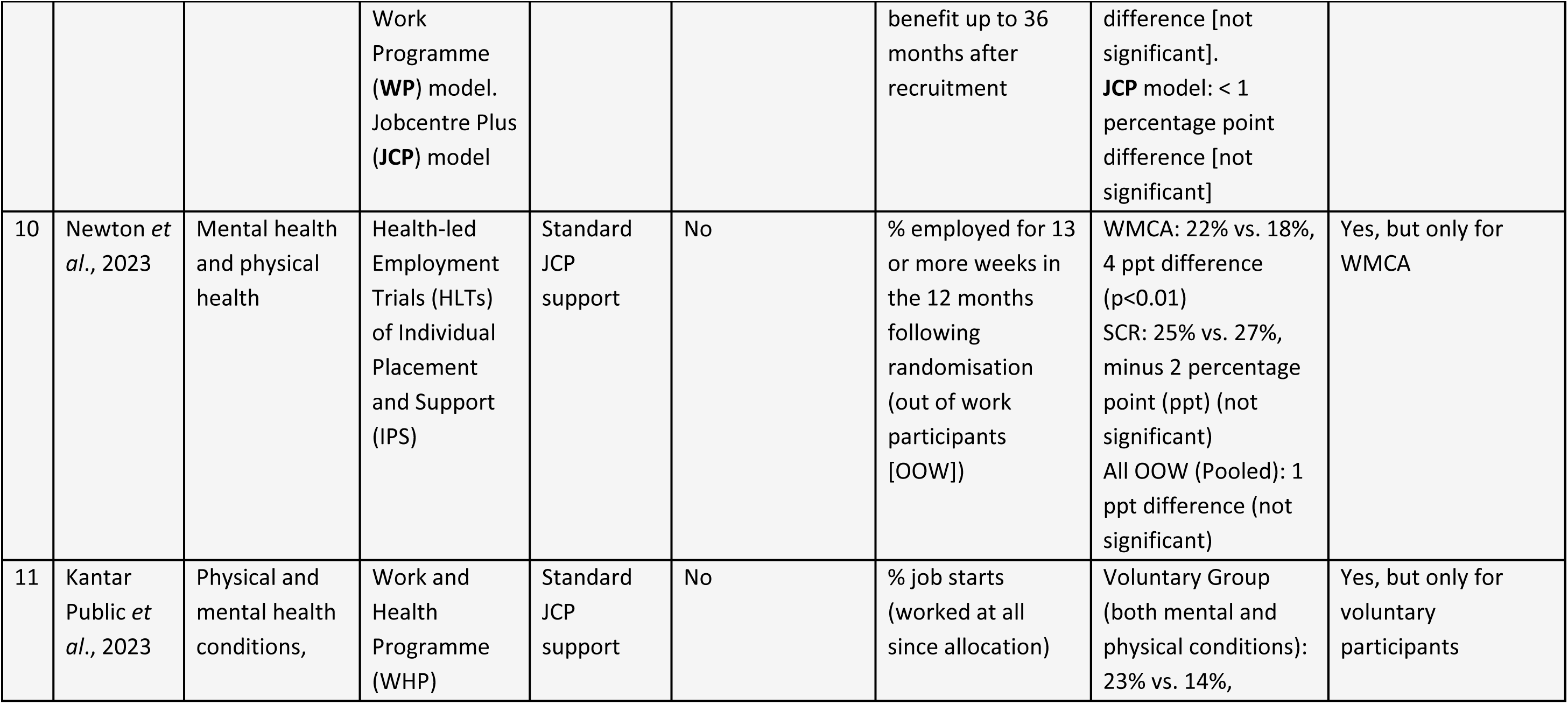
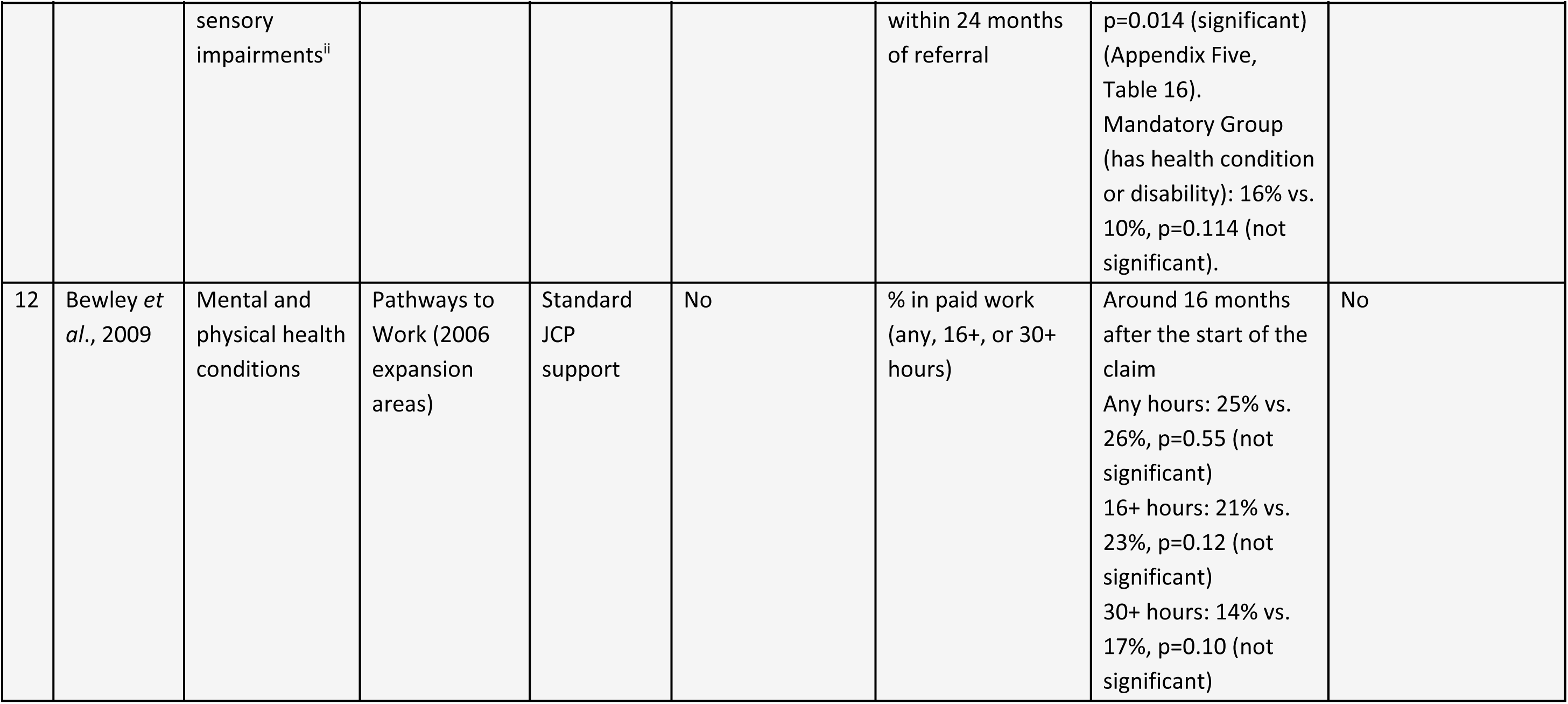
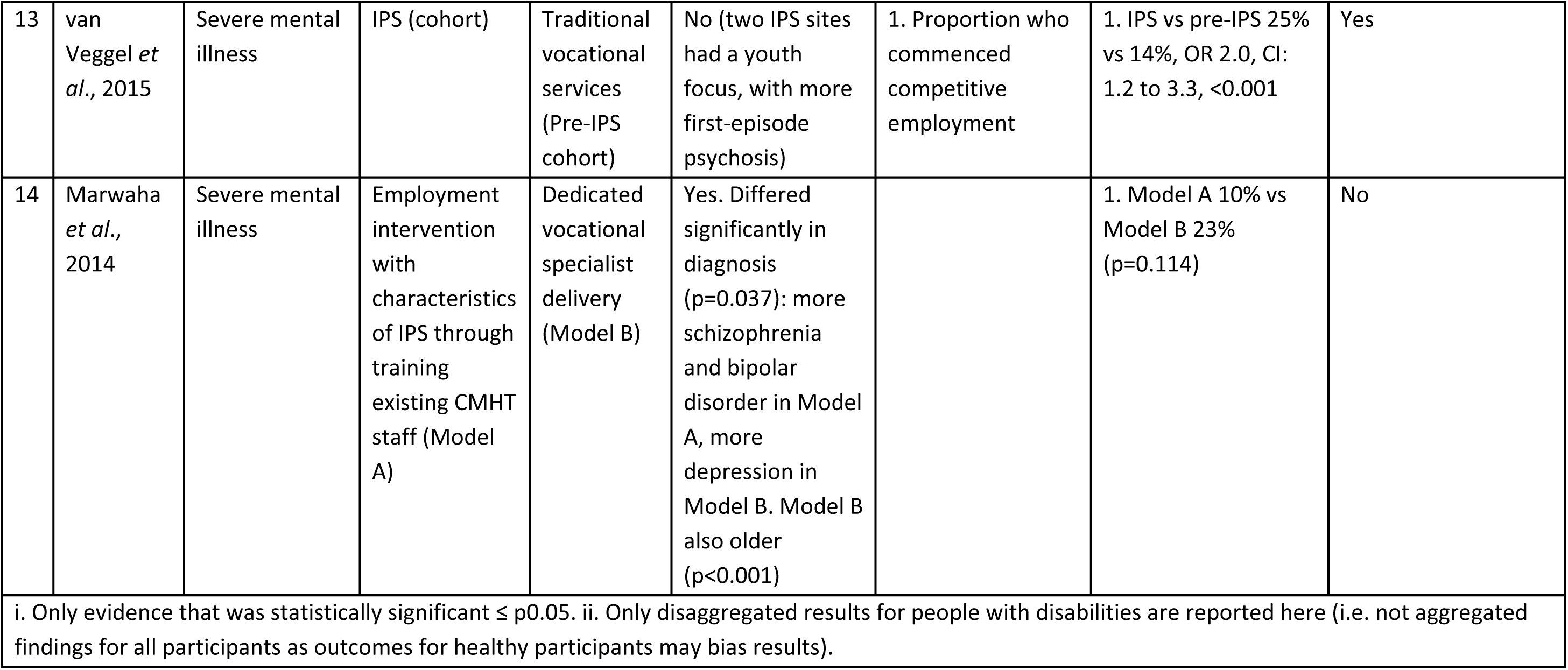
Study findings.

| N o. | Article | Population health condition/s | Intervention group (INT) | Control/co mparison group (COM) | Significant demographic differences between intervention & control groups at baseline (Y/N)? | Outcome description | Result – <u>Intervention</u> compared to control group (see notes i, ii) | Any evidence<sup>i</sup> intervention was more effective than control for employment rates? |
| --- | --- | --- | --- | --- | --- | --- | --- | --- |
| 1 | Craig *et al.*, 2014 | Early psychosis | IPS & motivational interviewing | IPS only | Yes, but adjusted in analysis | 1. % in open employment at 12 months | 1. OR = 4.3, 95% CI: 1.5 to 16.6 | Yes |
| 2 | Heslin *et al.*, 2011 | Severe mental illness | IPS | Traditional vocational services | No | % in open competitive employment | 22% vs. 11%, adjusted p=0.041 (unadjusted p=0.053) (significant after adjustment) | Yes |
| 3 | Moran, 2017 | Mental health, physical health, substance use disorders | MIS (RCT)<br>VEI (PSM) | Standard JCP support (MIS trial)<br>Propensity Score Matching (VEI ) | No | MIS trial: % in employment (monitored at weekly intervals)<br>Reduction in time spent on DWP benefit<br>VEI trial: 1. % in employment<br>Reduction in time spent on DWP benefits | MIS trial: employment difference not statistically significant (cumulative 0.5 ± 3 days in employment); 3.2 fewer days on benefit<br>VEI trial: just states “no measurable impact (no data given) | No |
| 4 | Howard *et al.*, 2010 | Severe mental illness | IPS | Traditional vocational services | No | % in open competitive employment | RR 1.35, 95% CI 0.95 to 1.93, p=0.15) [not significant] | No |
| 5 | Burns *et al.*, 2015 | Severe mental illness | IPS-LITE (time limited IPS) | IPS only | No | Competitive employment:<br>1. Worked for at least 1 day<br>2. Worked for at least 13 weeks | 1. OR = 0.82 (CI: 0.38 to 1.78) [not significant]<br>2. OR = 0.75 (CI: 0.32 to 1.73) [not significant] | No |
| 6 | Lindley *et al.* 2015 | Mixed (Incapacity Benefit recipients) | Want2Work scheme | Labour Force Survey job-seekers outside pilot areas (statistical control) | Yes, but matched in analysis | % gaining employment within 15 months | Matched estimate: 7 to 8 percentage points more likely to find employment (raw: 30% vs 19% of all out-of-work LFS respondents) | Yes |

| N o. | Article | Population health condition/s | Intervention group (INT) | Control/comparison group (COM) | Significant demographic differences between intervention & control groups at baseline (Y/N)? | Outcome description | Result – <u>Intervention</u> compared to control group (see notes i, ii) | Any evidence<sup>i</sup> intervention was more effective than control for employment rates? |
| --- | --- | --- | --- | --- | --- | --- | --- | --- |
| 7 | Major *et al.*, 2010 | First episode psychosis | Modified IPS programme (greater emphasis on education) | Usual care (early intervention service) | No | % in competitive employment at any time during 12 months | 36% vs 19% (no significance test reported). Context: adjusted OR for vocational recovery (employment or education) 3.53, CI: 1.25 to 10.00 | Not tested |
| 8 | Adams *et al.*, 2022 | Mental and physical health conditions, and learning difficulties | Integrated psychological therapy and employment support | Psychological therapy only | No | % employed within 12 months | 48% vs. 29%, p=0.015 (clients looking for work) | Yes |
| 9 | Dawson *et al.*, 2019 | Mental and physical health conditions | Enhanced JCP support: Healthcare Provider (HCP) model. | Standard JCP support | No | Employment rates (derived from HMRC P45 start and end dates). Accumulated additional days off | **HCP** model: < 1 percentage point difference [not significant].<br>**WP** model: < 1 percentage point | No |
|  |  |  | Work Programme (WP) model. Jobcentre Plus (JCP) model |  |  | benefit up to 36 months after recruitment | difference [not significant]. JCP model: < 1 percentage point difference [not significant] |  |
| 10 | Newton *et al.*, 2023 | Mental health and physical health | Health-led Employment Trials (HLTs) of Individual Placement and Support (IPS) | Standard JCP support | No | % employed for 13 or more weeks in the 12 months following randomisation (out of work participants [OOW]) | WMCA: 22% vs. 18%, 4 ppt difference (p<0.01)<br>SCR: 25% vs. 27%, minus 2 percentage point (ppt) (not significant)<br>All OOW (Pooled): 1 ppt difference (not significant) | Yes, but only for WMCA |
| 11 | Kantar Public *et al.*, 2023 | Physical and mental health conditions, | Work and Health Programme (WHP) | Standard JCP support | No | % job starts (worked at all since allocation) | Voluntary Group (both mental and physical conditions): 23% vs. 14%, | Yes, but only for voluntary participants |

| N o. | Article | Population health condition/s | Intervention group (INT) | Control/comparison group (COM) | Significant demographic differences between intervention & control groups at baseline (Y/N)? | Outcome description | Result – <u>Intervention</u> compared to control group (see notes i, ii) | Any evidence<sup>i</sup> intervention was more effective than control for employment rates? |
| --- | --- | --- | --- | --- | --- | --- | --- | --- |
|  |  | sensory impairments<sup>ii</sup> |  |  |  | within 24 months of referral | p=0.014 (significant) (Appendix Five, Table 16).<br>Mandatory Group (has health condition or disability): 16% vs. 10%, p=0.114 (not significant). |  |
| 12 | Bewley *et al.*, 2009 | Mental and physical health conditions | Pathways to Work (2006 expansion areas) | Standard JCP support | No | % in paid work (any, 16+, or 30+ hours) | Around 16 months after the start of the claim<br>Any hours: 25% vs. 26%, p=0.55 (not significant)<br>16+ hours: 21% vs. 23%, p=0.12 (not significant)<br>30+ hours: 14% vs. 17%, p=0.10 (not significant) | No |
| 13 | van Veggel *et al.*, 2015 | Severe mental illness | IPS (cohort) | Traditional vocational services (Pre-IPS cohort) | No (two IPS sites had a youth focus, with more first-episode psychosis) | 1. Proportion who commenced competitive employment | 1. IPS vs pre-IPS 25% vs 14%, OR 2.0, CI: 1.2 to 3.3, <0.001 | Yes |
| 14 | Marwaha *et al.*, 2014 | Severe mental illness | Employment intervention with characteristics of IPS through training existing CMHT staff (Model A) | Dedicated vocational specialist delivery (Model B) | Yes. Differed significantly in diagnosis (p=0.037): more schizophrenia and bipolar disorder in Model A, more depression in Model B. Model B also older (p<0.001) |  | 1. Model A 10% vs Model B 23% (p=0.114) | No |
| i. Only evidence that was statistically significant $\leq p0.05$. ii. Only disaggregated results for people with disabilities are reported here (i.e. not aggregated findings for all participants as outcomes for healthy participants may bias results). | | | | | | | | |

Findings on IPS trials are reported first below, followed by findings from evaluations of government employment programmes or pilots (trials). Higher methodological quality studies are reported first for each group of studies.

#### IPS interventions

Three articles from two studies rated as **very high** methodological quality evaluated IPS interventions for participants with severe mental health conditions.

In an RCT of an IPS intervention in South London, Howard *et al*. (2010). found that differences between an IPS intervention group and a control group in finding competitive employment were not statistically significant at 1-year follow up (RR 1.35, 95% CI 0.95–1.93, p=0.15). In a later two-year follow up, Heslin *et al*. (2011) found that a higher proportion of the IPS intervention group members found competitive employment compared to the control group receiving traditional vocational services. The proportion finding employment in both groups was, however, modest (22% vs. 11%; p=0.041 after adjustment for baseline characteristics, unadjusted p=0.053).

A cluster-RCT, compared a modified version of IPS that incorporated motivational interviewing to tackle negative preconceptions of barriers to employment amongst patients and early intervention team clinicians, with IPS only. Craig et al. (2014) found that the intervention group had significantly higher odds, more than fourfold, of being in competitive employment at 12 months than the control group receiving IPS only (OR=4.3, 95% CI: 1.5 to 16.6).

Two studies rated as **high** methodological quality assessed the effectiveness of modified forms of IPS for participants with severe mental illness.

In a prospective cohort study, Major *et al*. (2010) compared a modified IPS programme for first episode psychosis in London, with a greater emphasis on education due to the typically younger age of the cohort. 36% of the modified IPS group achieved competitive employment at some stage of a 12-month follow-up, compared to 19% in the comparison group, although no significance test was reported for this outcome. The authors’ primary outcome, ‘vocational recovery’ (competitive employment or education leading to a recognised qualification), was more likely with access to the intervention after adjustment (OR=3.53, 95% CI: 1.25 to 10.00; unadjusted OR=1.80, 95% CI: 0.84 to 3.88). A further 20% of the intervention group and 24% of the comparison group were in education.

An RCT comparing a modified, time limited, version of IPS (IPS-LITE) with IPS only found that rates of employment were similar at 18 months for IPS-LITE and non-time limited IPS (41% versus 46%, respectively). There were no statistically significant differences between the groups for other employment related outcomes (Burns et al., 2015).

Two studies of IPS for participants with severe mental illness were rated as **low** methodological quality in comparison to the other included studies.

Van Veggel *et al*. (2015) compared employment outcomes for an IPS cohort with a pre-IPS cohort and found the IPS cohort had twice the odds of commencing competitive employment (25% vs 14%, OR 2.0, CI: 1.2 to 3.3, p<0.001).

Marwaha *et al*. (2014) found that 23% of participants supported by a dedicated specialist delivering IPS within a CMHT (Model B) achieved an employment outcome, compared to 10% supported by CMHT staff who had received training to deliver a similar intervention (Model A). The result, however, was not statistically significant (p=0.114). Because the study was underpowered, with smaller than intended sample sizes, the researchers could only describe the 23% vs. 10% result as a "trend" suggesting Model B may be more effective, rather than statistically proven.

#### Government employment programmes or trials

One study (Moran, 2017) of the **2015 ESA trials** rated as very high methodological quality evaluated enhancements to Jobcentre Plus (JCP) support in parts of England, Scotland and Wales. They found no statistically significant differences in employment rate outcomes between either of the trial groups (More Intensive Support, and the Voluntary Early Intervention) and the standard JCP support control group

One study (Lindley *et al*. 2015) of the Want2Work scheme for people on Incapacity Benefit in Wales rated as high methodological quality found that 30% of participants gained employment, compared with 19% of the full Labour Force Survey out-of-work sample and 32% of the job-seeker comparison group. After propensity score matching, participants were about 7 to 8 percentage points more likely to find employment.

Five studies were rated as medium **methodological quality**.

Adams *et al*.’s, (2022) cluster-RCT of the **Employment Advisers in IAPT** programme found that receiving integrated employment support in addition to clinical therapy significantly increased employment rates within 12 months. Among clients looking for work at entry, nearly half (48%) of those who saw an Employment Adviser were employed 12 months later, compared to 29% of the matched comparison group who only received clinical therapy (p=0.015); the report describes these estimates as indicative because of small samples (n=223 vs 68).

Dawson *et al*.’s (2019) RCT of the **ESA Work-Related Activity Group pilots** providing enhanced JCP support found that none of the enhanced provider support models, delivered by either JCP Work Coaches, health professionals, or private contractors, led to significantly increased employment rates than standard JCP support. The authors note that the HMRC P45 data where employment rates were derived from excludes self-employment or low-threshold earners and misses 20-30% of employment spells (lower earnings being particularly likely among employment programme participants). They also found potentially contradictory evidence that benefit claimant rates decreased significantly.

Newton *et al*.’s (2023) RCT of **Health-led Employment Trials** of IPS for participants with low-to-moderate mental health and physical health conditions, found mixed results, compared to standard JCP support. The percentage employed for 13 or more weeks in the 12 months following randomisation was only significantly higher for participants in the West Midlands Combined Authority (WMCA) group compared to the control group (22% vs 18%, p < 0.01). Results for the Sheffield City Region group and both groups/areas pooled were not statistically significant.

In the disaggregated results of Kantar Public *et al*.’s (2023) RCT of the **Work and Health Programme**, only voluntary group participants with both mental and physical health conditions achieved a statistically significantly higher employment rate than the control group (23% vs 14%, p=0.014). The employment rate difference for mandatory intervention group participants with a health condition or disability was not statistically significant, compared to the control group (16% vs 10%, p=0.114).

Bewley *et al*.’s (2009) quasi-experimental study found no statistically significant evidence that the **Pathways to Work programme** (intervention group) increased the proportion of participants in paid work (Any hours: 25% vs. 26%, p=0.55; 16+ hours: 21% vs. 23%, p=0.12; 30+ hours: 14% vs. 17%, p=0.10). Subgroup results by gender, age, type of health condition (mental vs. physical), or having dependent children were also not statistically significant.

### Education and work history profile of intervention participants

The table in Appendix 8 provides a comparison between the educational attainment and previous employment history of intervention participants and the average for the UK population who are economically inactive due to long term sickness or disability. This group UK population group are very disadvantaged and experience low levels of labour market attachment. In 2019 only 8% have degree level qualifications and 37% have no qualifications. Only 9% have worked in the past 2 years and 23% have never worked (see Appendix 8 for details). For a large proportion of the studies (n=6, 50%) – the participants in the intervention evaluated were markedly better educated and had greater labour market attachment. For example in Adams et al.’s, (2022) cluster-RCT of the Employment Advisers in IAPT programme that found large effects on employment – 40% of participants looking for work had degree-level (Level 5+) qualifications and only 6% had never worked. Its is likely therefore that these results would not be generalisable to the bulk of people economically inactive due to ill health, many of whom will tend to be much further form the labour market. Overall studies of IPS were more likely to include participants who were less disadvantaged than the average, compared to non-IPS studies. Studies interventions evaluated as effective were more likely to include participants that were less disadvantaged than the average (see Appendix 8)

## Discussion and Conclusions

### Strengths of the review

The strict inclusion and exclusion criteria were designed to maximise internal and (contextual) external validity of the evidence to optimise potential relevance/transferability of findings to UK settings. Studies with control or comparison groups are inherently methodologically stronger forms of study design that reduce potential influence of confounders and help to account for underlying trends such as changes in employment markets that may have affected the outcomes at the same time as the interventions. The findings are more likely to be generalisable to the UK than previous reviews covering wider international geographies/contexts, given the similarities in social, cultural, economic, health, employment, and policy contexts of the included studies. It was also a strength to include studies of interventions where RCTs were not practical, and also using routinely collected data across large population groups in studies such as Lindley *et al*.’s (2015) study of the Want2Work scheme in Wales. Assessing the educational and work history profile of participants in evaluated interventions enables a better assessment of the generalisability of evidence to the overall population out of work with a disability in the UK.

### Limitations of the review

The strict inclusion criteria limited the range and coverage of evidence, including the types of employment interventions and potentially the health conditions of participants. The nature of IPS interventions may make them more suitable for RCT evaluations than other interventions. This may have led to a predominance of IPS RCT evaluations in the higher methodological quality evidence base. The location and inclusion of evidence from grey literature publications was made logistically possible through new hybrid human-AI approaches that increased effectiveness and efficiency of study location and data extraction processes. This allowed evaluations of a wider range of larger-scale government employment programmes/trials and population groups/health conditions to be included.

### Limitations and gaps in the evidence-base and areas for further research

There was a lack of high-quality evidence in the peer reviewed academic literature on the effectiveness of employment interventions for people with physical health conditions/disabilities, and multimorbidity. This is consistent with limitations in the international evidence base. The grey literature publications did, however, include evaluations of a broader range of mental and physical health conditions and disabilities.

The coverage of the evidence across the countries of the UK was limited. Study populations tend to be more advantaged, having higher qualifications and more recent employment history than the average for people out of work with disabilities in the UK. This is particularly true for smaller scale targeted interventions such as IPS. This may be due to selection of more advantaged groups into those interventions, or selection into trial enrolment. This limits the generalisability of evidence and its potential role in reducing the disability employment gap. There is an urgent need for evidence for the effectiveness of approaches for people with lower levels of education, who have limited labour market attachment. Greater geographical coverage of the UK evidence base, representing different health, social, economic, employment, and policy contexts in different nations and regions should be improved.

Table 3 helps to highlight methodological limitations across the studies, indicating issues with blinding and levels of participant dropout. Both these issues are, however, to be expected as it is difficult to conceal treatment groups for these types of interventions, and because study dropout rates may be influenced by the severity and nature of a participant’s health conditions. Information within some articles on participant selection and representativeness was also limited. It could be an indication of selection bias across some of the studies, or simply that recording and reporting should be improved.

Although the review sought to locate high methodological quality evidence on both micro- and meso-level employment interventions for people with disabilities in the UK, evidence that met the inclusion criteria was more focussed on micro-level interventions (individuals). High methodological quality UK evidence on environmental improvements by employers appears to be lacking. The nature of UK employment has changed in recent years, with new technologies, jobs and working practices having increasing impacts (e.g. assistive technologies, self-employment in platform economies, remote and hybrid working). The emerging body of evidence on recent meso-level approaches suggests they may be particularly beneficial for people with disabilities (e.g., Holland *et al*., 2026). That body of evidence should be strengthened and widened. Both the positive and negative impacts of recent changes to the nature of the "world of work" across population groups needs to be understood, and potential implications to design, delivery, and outcomes of employment interventions for people with disabilities addressed to help narrow the disability employment gap and tackle economic inactivity across the UK.

### Addressing transparency of government evaluation

A critical barrier to high-quality evidence in the UK is the transparency of government evaluations. Although The Magenta Book Guidance sets central standards for robust policy appraisal, major spending departments—particularly the Department for Work and Pensions (DWP)—have withheld or delayed publication of some commissioned evaluations and internal reviews, leaving findings in the grey literature (HM Treasury, 2026; House of Commons Work and Pensions Committee, 2022; Mills and Pring, 2024).

To reconcile political sensitivities with scientific rigour - and prevent ministers from delaying publication indefinitely - the UK should build on Treasury evaluation guidance by establishing a statutory ‘embargo-and-release’ protocol. Under this model, commissioned evaluations would be prepared for independent peer review but held under an administrative embargo, to be released only upon a change of government or after a maximum limit of five years. This structure insulates sitting administrations from immediate partisan scrutiny while guaranteeing that the wider scientific and policy communities gain eventual, unredacted access to vital evidence.

#### Implications for Policy: From High-Fidelity Foundations to Structural Support

The IPS (Individual Placement and Support) evidence indicates the need for high fidelity foundations along with testing modular enhancements. Although individual IPS (Individual Placement and Support) evaluations report varying absolute employment rates, reflecting differences in local labour markets and clinical populations - the *direction* of effect is consistent across the included trials: high-fidelity, specialist-led IPS tends to outperform traditional, non-integrated vocational services (Heslin et al., 2011). This consistency suggests that supported employment’s effectiveness is a structural property of the model itself that may be generalisable to diverse populations. We therefore propose a simple hierarchy for policy: a high-fidelity baseline, augmented by targeted modular enhancements.

The foundation should be dedicated clinically integrated specialists. The evidence suggests that diluting the core IPS model reduces its effectiveness. Marwaha et al. (2014) found that training existing, generalist Community Mental Health Team (CMHT) staff to deliver employment support (Model A) achieved less than half the job-entry rate of dedicated specialist delivery (10% vs. 23%; p = 0.114, non-significant) - generalist case managers, juggling competing clinical duties, appear structurally unable to sustain the intensive, rapid job-search support IPS requires. Co-locating dedicated Employment Specialists - closely adherent to core IPS principles and tightly integrated within clinical multidisciplinary teams (GP surgeries, CMHTs, psychological therapy services) - should therefore be treated as the needed baseline of intervention design. The capacity of clinical teams to sustain this integration alongside their existing workload, and any knock-on effects on other services, urgently needs evaluation.

Once a high-fidelity foundation is secure, three evaluated adaptations suggest further gains are possible, each addressing a specific barrier:

##### Behavioural (motivational interviewing)

Where negative self-perception or clinical/institutional ambivalence bottlenecks job-seeking, adding Motivational Interviewing to standard IPS was associated with more than four times the odds of competitive employment (OR 4.3, 95% CI 1.5–16.6; Craig et al., 2014).

##### Educational (supported education)

For younger cohorts whose vocational development was interrupted by acute illness - e.g., first-episode psychosis - adding a supported-education component was associated with higher competitive employment (36% vs 19%, not tested for significance) and over three times the odds of vocational recovery, a combined employment and education outcome (adjusted OR 3.53, 95% CI 1.25–10.00; Major et al., 2010), potentially bridging clinical recovery and long-term educational attainment.

##### Structural (time-limited support)

To address service capacity constraints, Burns et al. (2015) tested a time-limited IPS model (IPS-LITE) and found no significant loss of long-term employment effectiveness compared with open-ended IPS (OR 0.82, 95% CI 0.38–1.78, ns) - a potential route to expanding throughput without diluting outcomes.

These enhancements were each tested in a single local context (Craig et al. in South London and the Midlands; Major et al. in Camden and Islington; Burns et al. in Oxford), so they cannot yet be assumed to generalise nationally. Before being codified into policy, they should be replicated across diverse labour markets and socioeconomic contexts, and tested in groups currently under-represented in the evidence base - those with a wider range of health conditions (e.g., common mental health problems, musculoskeletal conditions) and those with low education and weak labour market attachment.

Evaluation of large UK government employment programmes indicate that programmes offering integrated, voluntary, and personalised support tend to show significant positive effects on employment, while programmes built around standard or enhanced generic Jobcentre Plus (JCP) casework, particularly where participation is mandatory, largely do not. Kantar Public et al.’s evaluation of the Work and Health Programme provides a within-study comparison: voluntary participants with both mental and physical health conditions achieved significantly higher employment than controls while mandatory participants with a health condition or disability did not. Both Moran (2017) and Dawson et al. (2019) evaluations of JCP enhancements and Bewley et al. (2009) evaluation of work focused interviews in pathways to work found no evidence of effect, whilst Want2Work (Lindley et al., 2015) combined voluntary participation, community-based advisers integrated with health professionals appeared to be effective.

The move toward localised employment support under the *Get Britain Working* strategy (Department for Work and Pensions et al., 2024) is a structured opportunity to deploy devolved Supported Employment frameworks. Channelling funding directly to English local authorities and Established Mayoral Strategic Authorities (e.g., Liverpool City Region, West Midlands, Greater Manchester) via Integrated Settlements could support genuinely place-based delivery (Green et al., 2024). These models should adopt the "place, train, and maintain" framework with disciplined caseload limits (maximum active caseloads per case manager averaging 20 under the Supported Employment Quality Framework, 25 under IPS; Department for Work and Pensions, 2026a) and integrate closely with local healthcare networks such as the WorkWell pilots, so that clinical and vocational support are synchronised.

Beyond intervention design, the evidence points to structural barriers that no individual-level model can resolve on its own. Addressing these requires action on the employer side, on delivery infrastructure, and on the legal and administrative architecture around employment support.

Policy, delivery and the evidence base are heavily weighted toward "fixing the individual," with comparatively little attention to the immediate working environment. Meso-level, employer-focused adaptations - assistive software, physical workplace modifications, remote/hybrid working technology - may be highly effective in preventing premature labour market exit, but remain under-evaluated. Similarly legal and welfare architecture, such as the *Right to Try Guarantee* (Department for Work and Pensions, 2025a) could reduce the risk of trying work and the *Employment Rights Act 2025 - requiring employers to refuse flexible working requests only where reasonable, removing the waiting days and lower earnings limit for statutory sick pay, and cutting the qualifying period for unfair dismissal from two years to six months* - could help prevent premature labour market exit driven by fluctuating health conditions (Parliament of the United Kingdom, 2025).Their impact on employment, health, and wellbeing outcomes should be rigorously tested alongside individual-focused interventions.

### Conclusions

Our review indicates three connected findings. Embedding employment specialists within clinical teams, primary care surgeries, psychological therapy services, and augmenting their work with behavioural techniques such as motivational interviewing likely increases employment entry compared with standalone vocational support. Alongside this, participation should be voluntary rather than mandatory: mandatory frameworks fail to deliver employment gains and appear to deter engagement. Voluntary, client-led pathways build trust, reduce the internalised stigma of worklessness, and achieve the greatest additionality for those facing the most severe socioeconomic disadvantage. Realising both of these findings in practice depends on the evidence base itself changing. It remains heavily skewed toward interventions that improve individual employability, with too little attention paid to what works for the most disadvantaged claimants or for changing the employment environment itself. Future research should shift toward evaluating employment support for those furthest from the labour market as well as employer-side, meso-level interventions such as enforcing flexible-working compliance and enabling remote or hybrid working technologies to prevent premature labour market exit among low-skilled workers. This should use robust counterfactual designs (RCTs, difference-in-differences, propensity score matching), 24–36-month longitudinal follow-up, and administrative tax data triangulated with self-report to resolve earnings blind spots. Building this rigorous, context-sensitive evidence base is essential to identifying "what works" across complex, place-based settings - and, ultimately, to improving employment and population health and wellbeing.

## Data Availability

All data in this systematic review is available in the original included publications.

## CRediT Authorship Contribution Statement

Andy Pennington: Conceptualisation, Methodology, Formal analysis, Investigation, Writing – original draft, Project administration. David Newbold: Data curation, Investigation, Validation, Writing – review & editing. Maximilian Limburg: Data curation, Investigation, Validation, Writing – review & editing. Mary Cronin: Data curation, Investigation, Validation, Writing – review & editing. Yang Yu: Data curation, Investigation, Validation, Writing – review & editing. Dorcas N. Magai: Validation, Writing – review & editing. Fatemeh Hoda Fallah: Validation, Writing – review & editing. Thomas Hampton: Validation, Writing – review & editing. Sue Jarvis: Conceptualisation, Local context analysis, Policy translation, Writing – review & editing. John White: Conceptualisation, Local context analysis, Policy translation, Writing – review & editing. Ben Barr: Conceptualisation, Methodology, Supervision, Funding acquisition, Writing – review & editing. Philip McHale: Conceptualisation, Methodology, Supervision, Funding acquisition, Writing – review & editing.

## Funding Acknowledgements

This research was commissioned by The Health Foundation, as part of the wider Economies for Healthier Lives research programme [Unique Award Reference Number FR-000003049]. It was supported by the UKRI grant Civic Health Equity: From Silos to Systems (CHESS) (ES/Z503022/1), and the NIHR Applied Research Collaboration North West Coast (NIHR200182). The views expressed are those of the authors and not necessarily those of the funding bodies, the NHS, the NIHR, or the Department of Health and Social Care.

## Appendices

## Appendix 1 UK disability employment interventions: systematic review search strategy

### Section A: Grey Literature Search Protocol & Web-Domain Targeting

Grey literature represents a primary source of welfare-to-work evaluation evidence in the UK. Because traditional academic indexes fail to capture these reports, we conducted exhaustive searches of government and organisational websites. Advanced Google searches were restricted to the UK web space and specifically targeted government web domains (.gov.uk) and specialist organisational portals (such as the Learning and Work Institute) using Boolean search operators:

- Targeted .gov.uk search syntax: site:gov.uk "employment support" and "disability" and ("evaluation" or "pilot" or "trial")
- Targeted organisational search syntax: site:learningandwork.org.uk "employment" and "health" and "evaluation"

To account for Google’s relevance prioritisation algorithms, manual screening of Google search results was systematically capped at the first 100 results for each query, as the relevance and additionality of subsequent hits deteriorated rapidly.

### Section B: AI-assisted search & saturation verification

To ensure that no key governmental pilots or third-sector evaluations were omitted, we supplemented domain searches with conversational AI platforms (ChatGPT 4o, Microsoft Copilot) acting as specialised search agents. This stage was designed to verify search saturation. Reviewers administered the following systematic prompt:

> *"Identify any publicly-funded government pilots, trials, or local authority programs in the UK (England, Wales, Scotland, Northern Ireland) published between 2009 and 2024 evaluating employment support or ‘welfare-to-work’ schemes for individuals with disabilities, limiting long-term illnesses, or mental health conditions. Specifically focus on evaluations containing quantitative control or comparison groups and list their corresponding DWP research report numbers, commissioning bodies, or publishing organisations."*

### Section C: Bibliographic database search strings

Upon identifying the core grey literature, the search strategy was translated into highly structured syntaxes for five gold-standard databases. The complete search string implemented in OVID MEDLINE is detailed below. Final searches across all databases were ran 17^th^ February 2026.

|  |  |
| --- | --- |
| 1 | (mental* OR disabled OR disabilit* OR chronic ill* OR chronically ill OR chronic sick* OR LLTI OR longstanding illness OR long-standing illness OR long-term sick* OR long-term ill* OR long-term condition* OR chronic condition* OR impair*).ti,ab. |
| 2 | (Work* OR job* OR vocation* OR labour OR labor OR unemploy* OR employment OR employab* OR inactiv*).ti,ab. |
| 3 | (welfare-to-work OR "welfare to work" OR "back to work" OR back-to-work OR "return to work" OR return-to-work OR "Households into work" OR "Working well" OR "Ways to work" |
|  | OR "Job Entry Targeted Support" OR JETS OR rehabilitat* OR training OR retraining OR advisor* OR counsellor* OR coach* OR access to work OR access-to-work OR pathways to work OR pathways-to-work OR support to work OR support-to-work OR Employ Me OR "Work and Health Programme").ti,ab. |
| **4** | (britain OR british OR england OR english OR scotland OR scottish OR wales OR welsh).mp. |
| **5** | exp Great Britain/ OR (uk OR gb OR united kingdom OR "great britain").mp. |
| **6** | 1 AND 2 AND 3 |
| **7** | 4 OR 5 |
| **8** | 6 AND 7 |
| **9** | (geriatric* OR infant OR neonat*).ti,ab. |
| **10** | (Afghanistan OR Albania OR Algeria OR Andorra OR Angola OR Antigua OR Argentina OR Armenia OR Austria OR Azerbaijan OR Bahrain OR Bangladesh OR Barbados OR Belarus OR Belgium OR Belize OR Benin OR Bhutan OR Bolivia OR Bosnia OR Botswana OR Brazil OR Brunei OR Bulgaria OR Burkina Faso OR Burundi OR Cabo Verde OR Cambodia OR Cameroon OR Canada OR Central African Republic OR Chile OR China OR Colombia OR Comoros OR Congo OR Costa Rica OR Ivoire OR Croatia OR Cuba OR Cyprus OR Czech Republic OR Denmark OR Djibouti OR Dominica OR Dominican Republic OR Congo OR Ecuador OR Egypt OR El Salvador OR Equatorial Guinea OR Eritrea OR Estonia OR Eswatini OR Ethiopia OR Faeroe Islands OR Finland OR France OR French Guiana OR Gabon OR Gambia OR Georgia OR Germany OR Ghana OR Greece OR Grenada OR Guatemala OR Guinea OR Guinea-Bissau OR Guyana OR Haiti OR Holy See OR Honduras OR Hong Kong OR Hungary OR Iceland OR India OR Indonesia OR Iran OR Iraq OR Ireland OR Israel OR Italy OR Jamaica OR Japan OR Jordan OR Kazakhstan OR Kenya OR Kuwait OR Kyrgyzstan OR Laos OR Latvia OR Lebanon OR Lesotho OR Liberia OR Libya OR Liechtenstein OR Lithuania OR Luxembourg OR Macao OR Madagascar OR Malawi OR Malaysia OR Maldives OR Mali OR Malta OR Mauritania OR Mauritius OR Mayotte OR Mexico OR Moldova OR Monaco OR Mongolia OR Montenegro OR Morocco OR Mozambique OR Myanmar OR Namibia OR Nepal OR Netherlands OR Nicaragua OR Niger OR Nigeria OR North Korea OR North Macedonia OR Norway OR Oman OR Pakistan OR Panama OR Paraguay OR Peru OR Philippines OR Poland OR Portugal OR Qatar OR Romania OR Russia OR Rwanda OR Saint Helena OR Saint Kitts OR Saint Lucia OR Saint Vincent OR San Marino OR Sao Tome OR Saudi Arabia OR Senegal OR Serbia OR Seychelles OR Sierra Leone OR Singapore OR Slovakia OR Slovenia OR Somalia OR South Africa OR South Korea OR Sudan OR Spain OR Sri Lanka OR Palestine OR Suriname OR Sweden OR Switzerland OR Syria OR Taiwan OR Tajikistan OR Tanzania OR Thailand OR Bahamas OR Timor-Leste OR Togo OR Trinidad OR Tunisia OR Turkey OR Turkmenistan OR Uganda OR Ukraine OR United Arab Emirates OR UAE OR United States OR USA OR Uruguay OR Uzbekistan OR Venezuela OR Vietnam OR Western Sahara OR Yemen OR Zambia OR Zimbabwe).ti,ab. |
| **11** | 8 NOT 9 |
| **12** | 11 NOT 10 |
| **13** | Limit 12 to (English language and humans and yr="2009 - 2024") |

Search results were checked to ensure they had successfully located a series of sample papers.

### Section D: Citation snowballing and expert networks

To guard against inconsistencies in indexing and terminology across disparate disciplines, we conducted retrospective forward and backward citation tracking (citation snowballing) on all included publications. Forward tracking was automated utilising the Web of Science Core Collection, Research Rabbit AI mapping tools, and Google Scholar to identify papers citing our included studies. Backward tracking involved manual screening of the reference lists of all included articles. Finally, we contacted established networks of UK disability employment researchers and regional partners (such as Combined Authority leads and DWP regional advisors) to identify emerging evaluations and papers in press.

### Section E: Screening, deduplication, and AI-assisted extraction

Search results were deduplicated in EndNote X9 using a three-stage validation protocol (one automated stage, followed by two independent manual review sweeps). Deduplicated references were imported into EPPI-Reviewer 6 for independent dual-screening at the title/abstract and full-text stages. Due to the substantial word count and formatting heterogeneity of grey literature reports, we deployed a hybrid extraction workflow: reviewers utilised Gemini Notebook and PDF semantic querying software to execute standardised data queries across massive reports (frequently exceeding 100 pages/100,000 words), with every extracted data point independently audited and verified against the primary document by a second human reviewer to ensure completeness and prevent extraction errors.

## Appendix 2 Quality assessment tool for systematic reviews of effectiveness (based on Orton, *et al*, 2016; modified from Lorenc *et al*., 2013)

First author: Year:

The quality assessment tool contains six questions:

1. Selection bias
2. Study design
3. Confounders
4. Blinding
5. Data collection
6. Withdrawals and dropouts.

Each question can get an A (high), B (medium) or C (low) or D (lowest, for study design) quality rating, as per the tool below.

The guidelines for the specific questions are as follows.

| 1. Selection bias | Rating |
| --- | --- |
| Selected study sample very likely to represent population from target area AND 80 to 100% response at baseline | A |
| Selected study sample very likely to represent population from target area AND 60 to 79% response at baseline; OR Selected study sample somewhat likely to represent population from target area AND 80 to 100% response at baseline | B |
| <60% baseline response; OR Somewhat likely to represent population AND <80% response; OR Not likely to represent population OR representativeness NR/unclear; OR Response rate at baseline not reported (NR) or unclear | C |

| 2. Study design | Rating |
| --- | --- |
| Control group and pre and post longitudinal data OR random allocation | A |
| No control group and pre and post longitudinal data; OR<br>Control group and pre and post cross-sectional data AND no indication of major change in population | B |
| No control group and pre and post cross-sectional data; OR<br>Control group and pre and post cross-sectional data AND possibility of major change in population | C |
| Control group, post intervention only data | D |
| **Note:** 'longitudinal' = same individuals pre and post; 'cross-sectional' = different individuals. Where studies use mixed designs (e.g. presenting both cross-sectional and longitudinal data), give the highest grade applicable to the analyses actually reported. Where studies collect longitudinal data and report attrition rates, grade as longitudinal even if only cross-sectional analyses are reported. |  |

| 3. Confounders | Rating |
| --- | --- |
| Control group matched on key variables (at least two of: SES or relevant proxies (area or individual level), gender, age, ethnicity (individual level)) AND supporting data presented; OR<br>Outcomes adjusted for key variables (at least two of: gender, age, ethnicity, SES) using appropriate methods | A |
| Stated that control group matched or 'similar', but supporting data not presented | B |
| No matching or adjustment reported AND likely to be substantial differences between groups; OR no information on differences between intervention and control group; OR no control group | C |
| **Note:** RCTs will be rated 'B' if no information on between-group differences is presented. |  |

| 4. Blinding | Rating |
| --- | --- |
| Both outcome assessors AND participants blind to allocation | A |
| Either outcome assessors OR participants blind to allocation | B |
| Blinding NR; OR no control group | C |

| 5. Data collection | Rating |
| --- | --- |
| Piloting or pre-testing of tool; OR checks on validity of data (e.g. verification of a percentage of responses); OR tool shown to be reliable in relevant population | A |
| Data collection tool based on previous research, but no piloting or checking, and reliability not demonstrated | B |
| Data collection unclear; OR tools not piloted, checked or based on previous research | C |

| 6. Withdrawals and dropouts | Rating |
| --- | --- |
| Attrition <20% | A |
| Attrition 21%-40% | B |
| Attrition >40%; OR attrition NR; OR cross-sectional data only | C |
| **Note:** Attrition is measured as the percentage of the baseline sample lost at final follow-up. |  |

## Appendix 3 Declarations of Interest statement

### Declaration of Interest

The authors declare that they have no known competing financial interests or personal relationships that could have appeared to influence the work reported in this paper.

### Funding

This research was commissioned by The Health Foundation, as part of the wider Economies for Healthier Lives research programme [Unique Award Reference Number FR-000003049]. It was supported by the UKRI grant Civic Health Equity: From Silos to Systems (CHESS) (ES/Z503022/1), and the NIHR Applied Research Collaboration North West Coast (NIHR200182). The views expressed are those of the authors and not necessarily those of the funding bodies, the NIHR, or the Department of Health and Social Care.

## Appendix 4 To be added to full peer reviewed publication

## Appendix 5 List of articles included in the review (in alphabetical order)

## Appendix 6 Future research agenda: methodological frameworks and evidence building in UK disability employment policy

### 1. Conceptual framing: control, stress, and workplace dynamics

Developing a robust, forward-looking research agenda for UK disability employment interventions requires drawing upon established theoretical concepts surrounding individual autonomy, workplace demand, and health inequalities. A foundational concept is the ‘demand-control-support’ model of workplace stress proposed by Karasek and Theorell (1990). This model posits that a combination of high psychological job demands and low decision latitude (control), particularly when combined with a lack of social support, acts as a primary stressor in the work environment (Karasek and Theorell, 1990; Van der Doef and Maes, 1999). Empirical evidence has long demonstrated that exposure to this high-demand, low-control stress profile is socioeconomically graded, occurring disproportionately among lower-skilled and manual occupational classes, and represents a critical mechanism explaining socioeconomic gradients in physical and mental health conditions like cardiovascular disease (Marmot et al., 1997; Kuper and Marmot, 2003).

This theoretical framework can be extended to the ‘living environment,’ defined as the households, communities, and societies in which individuals live outside of paid employment (Pennington et al., 2018). Here, ‘control over destiny’-or the freedom to exercise meaningful choices over daily life-is proposed as an essential determinant of health and wellbeing (Syme, 1989; Marmot, 2004; Sen, 2003). Longitudinal evidence confirms that lower socioeconomic positions are systematically associated with lower control beliefs (or perceived control) in the living space, and that these low control beliefs explain a substantial proportion of the association between social disadvantage and adverse outcomes such as heart disease, anxiety, and depression (Orton et al., 2019). However, a critical gap remains: while the psychological pathway of ‘perceived control’ is well-documented, very few studies have attempted to measure ‘actual control’-the degree of objective decision latitude individuals can exercise over material and environmental resources (Orton et al., 2019; Whitehead et al., 2016). A comprehensive research agenda for disability employment must therefore bridge individual-level support with the objective, place-based material conditions of targeted populations.

### 2. Evaluating the disability employment evidence base

A rigorous future research agenda must build upon and contrast with the historical reviews of UK disability employment policy. Bambra et al. (2009) evaluated organisational changes to the psychosocial work environment (such as job redesign and shift modifications), demonstrating positive impacts on employee wellbeing but noting that underlying primary studies lacked causally robust designs. Clayton et al. (2011) evaluated individual-focused UK return-to-work initiatives (e.g., Pathways to Work), concluding that personal adviser models were severely limited by output-driven, binary targets that pressurised private contractors into ‘cream-skimming’ work-ready claimants whilst neglecting those furthest from the labour market. Clayton et al. (2012) reviewed employer-focused initiatives, finding that workplace adjustments had positive impacts on employment but low uptake, and that some interventions favoured more advantaged disabled people and those closer to the labour market. Similarly, Vooijs et al. (2015) evaluated generic work-directed interventions across chronic diseases, finding that interventions directed at the work environment were mostly reported as effective. While Bond et al. (2023) demonstrated that the Individual Placement and Support (IPS) model was effective for young adults, their review was restricted to young adults with serious mental illness or early psychosis.

This review advances this literature by evaluating UK-specific interventional evidence (2009–2024) through the dual lenses of socioeconomic disadvantage and common mental health conditions (such as anxiety, depression, and stress). It demonstrates that high-fidelity IPS is robustly effective at helping UK individuals with severe mental health conditions into competitive employment, outperforming traditional vocational services (Heslin et al., 2011). Crucially, this review highlights that specific interventional modifications - such as combining IPS with motivational interviewing to resolve participant and advisor ambivalence-achieved a dramatic fourfold increase in competitive employment odds (Craig et al., 2014), while an educational emphasis was associated with vocational recovery for younger cohorts (Major et al., 2010). Furthermore, integrating Employment Advisers directly within clinical psychological therapy teams (IAPT) increased 12-month employment rates from 29% to 48% (Adams et al., 2022). In contrast, this review confirms that large-scale, mandatory government programmes (e.g., the 2015 ESA trials and the ESA Work-Related Activity Group pilots) yielded no statistically significant increases in competitive employment rates (Moran, 2017; Dawson et al., 2019), whereas voluntary, client-led participation models (such as Want2Work or the Work and Health Programme voluntary cohort) were associated with higher employment (Lindley et al., 2015; Kantar Public et al., 2023).

### 3. A multi-method research agenda to enhance methodological strength

#### 3.1 Tackling internal validity and primary evidence quality

To build the quality of both review-level and primary-level evidence over time, future research must systematically address threats to internal validity, such as selection bias, the lack of blinding, and high participant dropout rates.

##### Managing selection and attrition

Primary interventional evaluations of voluntary programs are highly susceptible to self-selection bias, where more motivated or healthier claimants enrol. Future trials should utilise robust counterfactual designs - such as randomised controlled trials (RCTs) or quasi-experimental methods like Difference-in-Differences (DiD) and Propensity Score Matching (PSM) - to ensure a reliable comparison group (Lindley et al., 2015; Adams et al., 2022). While clinical blinding of participants or coaches is rarely practical in vocational interventions, trials must utilise independent, blinded data analysts to prevent reporting bias.

##### Enhancing outcome measurement

Research must move away from short-term (6–12 month), binary ‘job starts’ which incentivise provider cream-skimming. Evaluators should track long-term longitudinal cohorts (24–36 months) to measure actual employment retention, wage trajectory, and health progression (Heslin et al., 2011). To eliminate cream-skimming, primary evaluations must develop and utilise standardised, validated instruments that measure intermediate progress or ‘movement toward work’ (such as confidence, self-efficacy, or CV creation) for those furthest from the labour market (Clayton et al., 2011).

##### Resolving administrative data blindspots

Evaluations of large-scale programs frequently rely on HM Revenue and Customs (HMRC) P45 tax data to monitor job entries. However, HMRC records under-represent self-employment and jobs with earnings below the tax threshold, and are likely to miss around 20% to 30% of employment spells (Dawson et al., 2019). Evaluators must combine administrative tax databases with self-reported earning logs and local trading records to prevent significant underestimation of program effects.

### 3.2 Enriching external validity and contextual realities

While maintaining internal validity, researchers must remain highly vigilant about external validity, ensuring that trial findings translate to real-world sociocultural and geographical contexts.

#### Accounting for place-based dynamics

Disability employment support is heavily moderated by local labour market conditions. For example, the Health-led Employment Trials of IPS (Newton et al., 2023) demonstrated a strong, statistically significant employment impact in the West Midlands Combined Authority region but showed no significant impact in the Sheffield City Region. Evaluators must systematically record and report regional economic indicators, housing markets, and transportation infrastructure to contextualise generalisability.

#### Aligning with contemporary claimant demographics

Primary evaluations must move beyond highly specific clinical cohorts (e.g., severe psychosis in Major et al., 2010) to reflect the modern profile of UK health-related welfare claimants. Mental and behavioural conditions account for around half of the post-pandemic rise in disability benefit claims (Latimer et al., 2025).

### 4. Specific overarching methodological approaches

#### 4.1 Theoretical grounding and logic models

Evaluators must co-produce explicit, theory-led logic models and pathways of change prior to interventional trials, mapping how individual-level (micro) modifications interact with organisational (meso) and local economic (macro) determinants of health inequalities (Whitehead et al., 2016; Orton et al., 2019). Figure 1 illustrates the iterative evidence-building cycle designed to continuously feed empirical outcomes back into theoretical conceptualisation.

**Figure 1:**
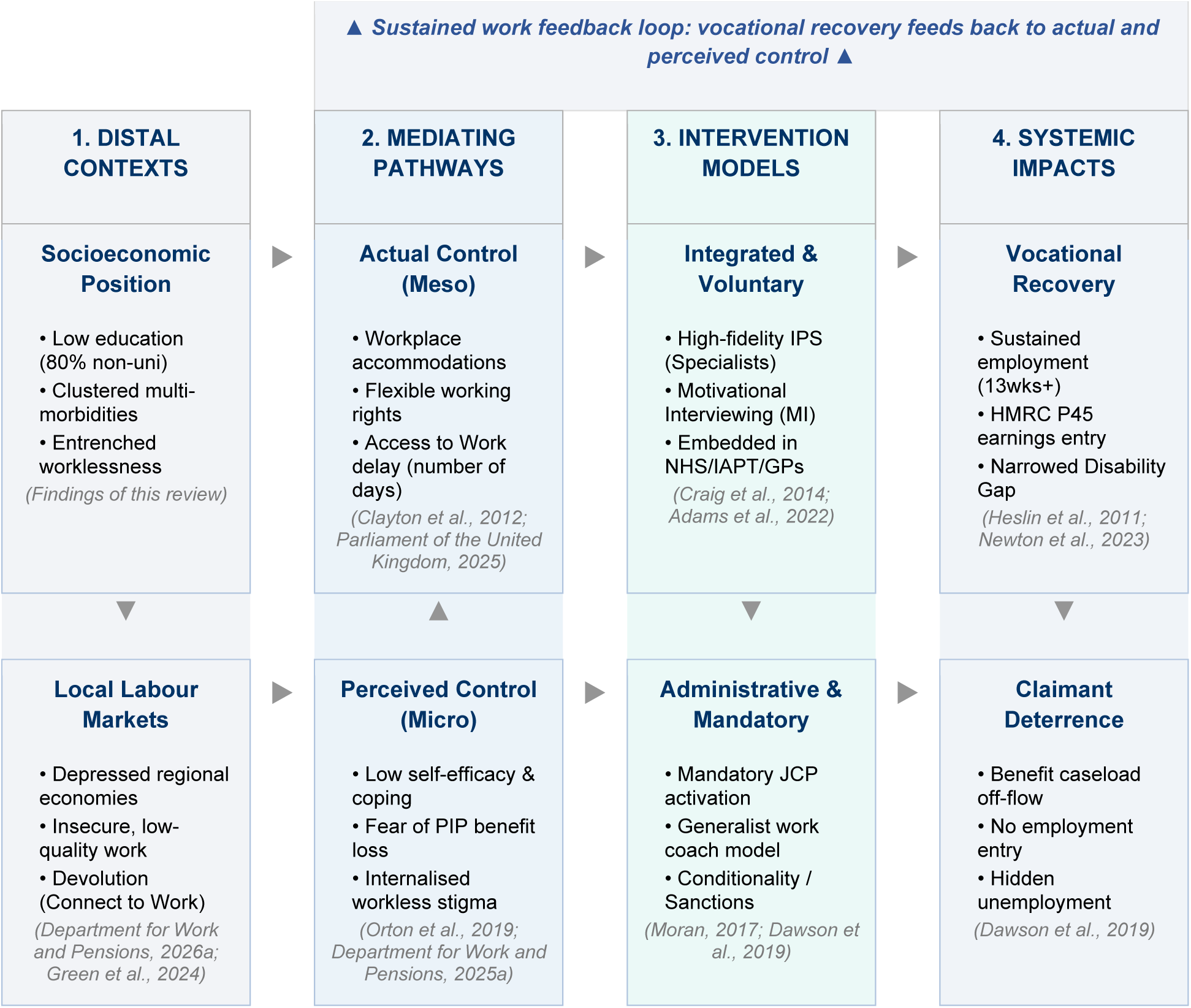
Model of the Iterative evidence-building cycle for devolved disability employment interventions.

#### 4.2 A Coordinated typology of evidence reviews

A robust research infrastructure requires a coordinated cycle of different review methodologies, drawing from both academic publications and grey literature sources:

##### Critical reviews of theories and concepts

Interrogate foundational concepts (e.g., Syme’s ‘control over destiny’ or Karasek’s ‘decision latitude’) to ensure primary interventions are targeted at modifiable structural barriers rather than unmodifiable personal traits (Orton et al., 2019).

##### Scoping reviews

Rapidly map the breadth and nature of emerging, less-developed literatures (e.g., self-employment in platform economies or the deployment of flexible remote-working technologies) to identify where evidence is sufficient for formal systematic reviews.

##### Systematic reviews

Synthesise evidence of interventional effectiveness with strict inclusion of control groups, providing a clear counterfactual to determine effect size in similar sociocultural and policy contexts.

##### Realist reviews / syntheses

Unpack the ‘what works, for whom, in what circumstances, and why’ (Pawson, 2006). Realist reviews are vital for complex, multi-component vocational programs because they focus on how local context triggers specific cognitive or behavioural mechanisms in participants, rather than treating the intervention as a simple linear cause.

##### Meta-reviews / umbrella reviews

Synthesise evidence across multiple pre-existing systematic reviews (e.g., Bambra et al., 2009; Vooijs et al., 2015) to identify high-level, generic principles of intervention effectiveness.

#### 4.3 Advancing mixed-methods integration

Complex social interventions cannot be fully understood through quantitative metrics alone. Future primary evaluations and reviews should utilise sequential exploratory or explanatory designs that combine:

##### Quantitative counterfactuals

Establish ‘if’ and ‘to what extent’ an intervention worked, using RCTs, DiD, or PSM to isolate the program effect from macroeconomic trends (Lindley et al., 2015; Adams et al., 2022).

##### Qualitative process evaluations

Explain ‘how’ and ‘why’ it worked (or failed), utilising semi-structured interviews, focus groups, and ethnographic observations of coaches, claimants, and employers. These explain implementation fidelity, claimant experiences, and context-mechanism-outcome (CMO) configurations (Craig et al., 2014; McGill et al., 2020).

##### Narrative and realist syntheses

Integrate these disparate data types textually, using narrative synthesis guidelines (Popay et al., 2006) to integrate quantitative effect sizes with qualitative process data, thereby telling the complete ‘story’ of the evidence.

#### 4.4 Equity-focused data collection and social determinants of health

To address health and wellbeing inequalities from a social determinants of health (SDH) perspective (Marmot et al., 2010), primary studies and reviews must systematically collect and disaggregate data using PROGRESS-Plus or other equity-focused frameworks. This ensures that evaluations can identify inequalities in program access, uptake, and effectiveness based on educational attainment, skill levels, age, gender, housing tenure, and Index of Multiple Deprivation (IMD) deciles (Adams et al., 2022).

### 5. Addressing transparency of government evaluation

The generation of high-quality UK evidence is hampered by political and administrative barriers. Thesection below details thecritical tension surrounding grey literature publication restrictions and proposes a novel institutional solution.

**TEXTBOX 1: Addressing the Grey Literature Publication Restriction in Government Evaluations**

**Tension:** UK government departments, including the Department for Work and Pensions (DWP), who commission most evaluations of large-scale publicly funded employment programs, frequently restrictor do not facilitate the publication of these evaluations in peer-reviewed academic journals (House of Commons Work and Pensions Committee, 2022; Mills and Pring, 2024). Consequently, these extensive evaluations remain trapped as unpublished report PDFs in the grey literature…"

**Reasons for Restriction:** The primary driver behind these restrictions is civil service risk a version and political control over the policy narrative (House of Commons Work and Pensions Committee, 2022; Mills and Pring, 2024). Active administrations fear that academic papers with nuanced, mixed, or statistically insignificant results will be politically weaponised by opposition parties or sensationalised by the media, potentially undermining the perceived success of flagship welfare-to-work policies.

**The embargo solution:** To reconcile political sensitivities with scientific rigor, government departments should establish a formal ‘embargo-and-release’ protocol. Commissioned evaluations should be prepared for academic peer-reviewed publication but placed under a strict statutory embargo until after a change of administration. This insulates the active administration from short-term partisan scrutiny, while ensuring that the wider scientific and policy communities eventually gain access to the highest-quality, peer-reviewed evidence to learn from previous policy cycles.

## Appendix 7 PRISMA 2020 and PRISMA-Equity Compliance Checklists

This appendix presents the complete reporting compliance checklists for the systematic review. Section A contains the PRISMA 2020 checklist, which maps the 27 items required for a standard systematic review. Section B contains the PRISMA-Equity extension checklist, which demonstrates how the review systematically addresses socioeconomic disadvantage, clinical conditions, and place-based labour market dynamics from a health equity perspective. Both checklists demonstrate that the manuscript aligns with the rigorous reporting standards of Social Science & Medicine.

Section A: PRISMA 2020 Checklist
| Item # | Reporting Standard Description | Compliance Status | Location / Compliance Details in Paper |
| --- | --- | --- | --- |
| 1 | Title: Identify report as systematic review. | FULLY MET | Title Page (p. 1) - clearly identified as a systematic review in the UK context. |
| 2 | Abstract: Provide a structured/unstructured summary. | FULLY MET | Abstract (p. 3) - highly compliant 205-word narrative summary meeting target journal requirements. |
| 3 & 4 | Intro: Describe rationale and specify objectives. | FULLY MET | Background & Objectives (p. 4-6) - fully mapped to the newly refined RQ1 and RQ2 questions. |
| 5 | Eligibility: Specify inclusion and exclusion criteria. | FULLY MET | Table 1 (p. 7) - comprehensively details all eligibility and exclusion criteria. |
| 6 | Info Sources: Specify databases, websites, and last search dates. | FULLY MET | Methods - Searches (p. 8) - explicitly states the databases used and the final search date: 17th February 2026. |
| 7 | Search Strategy: Present full replicable search strategy. | FULLY MET | Appendix 1 - fully details the OVID MEDLINE search strategy and search strings. |
| 8 | Selection Process: State independent dual screening methods. | FULLY MET | Methods - Screening (p. 9) - independent screening by two reviewers in EPPI-Reviewer 6, with manual dual screening of grey literature. |
| 9 | Data Extraction: State methods used to collect data, including AI. | FULLY MET | Methods - Data extraction (p. 9) - dual extraction, checked, AI assistance (Gemini Notebook) reported under strict human audit. |
| 10 | Data Items: List and define all variables sought. | **FULLY MET** | Methods - Data extraction (p. 9) - details setting, health conditions, predictors, and employment outcomes sought. |
| 11 | Quality Appraisal: Describe risk of bias/quality assessment tool. | **FULLY MET** | Methods - Methodological quality assessment (p. 10) - modified Orton et al. tool fully documented in Appendix 2. |
| 12 | Effect Measures: Specify effect metrics evaluated. | **FULLY MET** | Methods - Data synthesis (p. 10) - explicitly defines primary effect measures (OR, RR, DiD coefficients, percentage point differences). |
| 13 | Synthesis: Describe narrative synthesis and decisions to pool. | **FULLY MET** | Methods - Data synthesis (p. 10) - narrative synthesis well-justified over meta-analysis due to clinical and interventional heterogeneity. |
| 14 | Reporting Bias: Describe methods to assess/minimise bias. | **FULLY MET** | Methods (p. 10) - contains our custom paragraph methodologically justifying grey literature searching to minimise publication bias. |
| 24a | Registration: State registry and registration number. | **FULLY MET** | Methods - Intro (p. 8) - includes the official PROSPERO registration ID: CRD42018103930. |
| 26 | Funding/COI: Competing interest declarations. | **FULLY MET** | Appendix 3 - formal Declarations of Interest and Funding statements fully integrated. |

Section B: PRISMA-Equity Extension Checklist
| PRISMA-E Item | PRISMA-E Guideline & Equity Extension Description | How Met in Paper |
| --- | --- | --- |
| **Title 1** | Identify the report as a systematic review focused on health equity (or including an equity-focused question/analysis). | Title Page (p. 1) - clearly identified as a systematic review in the UK. The subtitle and main text explicitly focus on addressing socioeconomic disadvantage and common mental health conditions. |
| **Rationale 3** | Describe the health inequity of interest, why it is unfair and avoidable, and how the intervention is expected to affect it. | Background & Objectives (p. 4-6) - defines the Disability Employment Gap (30 percentage points) and how poorer health and multimorbidities cluster among low-skilled, low-education individuals in depressed regional labour markets. |
| **Protocol 5** | State if equity was explicitly considered in the review protocol | Methods - Intro (p. 8) - includes PROSPERO registration (CRD42018103930), which pre- |
|  | and whether the protocol was registered with equity as a priori focus. | specified socioeconomic disadvantage and mental health conditions as a priori equity focus areas. |
| **Eligibility 6** | Describe the population characteristics used to define equity groups (e.g., PROGRESS-Plus elements like Education, Occupation, Place, Social Capital) and the justification for these boundaries. | Methods - Inclusion/Exclusion (Table 1) - specifically targets individuals with disabilities, and PROGRESS-Plus disadvantage is defined using low qualifications/skills, long-term worklessness, and geographic residency in weak local labour markets. |
| **Information Sources 7** | Describe the search strategy for equity-relevant studies, including grey literature domains and supplemental approaches. | Methods - Searches (p. 8) - describes our exhaustive grey literature search protocol (including .gov.uk and Learning and Work Institute, hand searches and extensive citation searches) to capture government evaluations, bypassing academic indexing biases. |
| **Search 8** | Provide the full search string used for at least one database, demonstrating how equity terms were incorporated. | Appendix 1 - contains the complete 13-line OVID MEDLINE search string, which incorporates search concepts for health conditions, employment, and UK geographical terms. |
| **Study Selection 9** | Describe the process of screening and selecting studies, including whether dual independent screening was applied. | Methods - Screening (p. 9) - screening managed in EPPI-Reviewer with dual independent screening at all stages, plus manual dual screening of the first 100 Google results for grey literature. |
| **Data Items 11** | List and define all variables for which data were sought, particularly those used to disaggregate or capture equity dimensions. | Methods - Data extraction (p. 9) - extracted variables include: population health conditions, measures of disadvantage, and geographical delivery settings. |
| **Risk of Bias 12** | Describe how risk of bias assessment accounted for equity aspects (e.g., representativeness, selection bias, attrition related to health status). | Methods - Methodological quality assessment (p. 10) - critical quality appraisal utilised a modified version of the Lorenc et al. (2013) tool, scoring studies across selection bias, representativeness, confounder adjustment, data collection, and attrition. |
| **Synthesis 14** | Describe how equity characteristics were synthesised, including disaggregated analysis or subgroup synthesis. | Methods - Data synthesis (p. 10) - narrative synthesis is structured around interventional typologies (high-fidelity IPS |
|  |  | vs. government programmes) and disaggregated outcomes. |
| **Results: Study Selection 17** | Provide a flow diagram showing the flow of studies, noting if any were excluded specifically due to lacking equity-focused disaggregation. | Results (p. 10) - PRISMA flow chart (Figure 1) details the flow of 8,088 records down to 14 included articles. Appendix 4 documents reasons for exclusion, such as pooling disabled populations with healthy workers. |
| **Results: Characteristics 18** | Present a table of characteristics for each included study, incorporating disaggregated equity data. | Table 2 ('Characteristics of included studies') includes dedicated, fully populated columns for 'Population health condition/s' and 'Measures of disadvantage' for all 14 included papers. |
| **Results: Outcomes 21** | Present results disaggregated by equity characteristics to show differential effectiveness across groups. | Table 4 ('Study findings') displays disaggregated results, specifically isolating outcomes for voluntary vs. mandatory cohorts, different local labour markets, and integrated vs. therapy-only groups. |
| **Discussion: Limitations 25** | Discuss study-level and review-level limitations with an equity lens. | Discussion - Limitations and gaps (p. 20) - identifies critical equity limitations: reliance on HMRC P45 tax data in government trials (excluding self-employment and low earners) and the severe gap in physical disabilities and multimorbidity. |
| **Discussion: Conclusions 26** | Provide policy implications that address equity, focusing on how interventions can narrow the gap for the most disadvantaged. | Discussion - Policy implications (p. 21) - outlines actionable, equity-focused recommendations, including the shift to voluntary Supported Employment, place-based targeting, and employer-side adjustments. |
| **Funding 27** | State sources of funding for the systematic review and describe any potential conflicts of interest. | Appendix 3 - formal Declarations and Funding statements integrated, citing the Health Foundation, UKRI, and NIHR Applied Research Collaboration. |

## Appendix 8 Comparison of Educational attainment and previous employment history for intervention-arm participants across the studies vs. the UK long-term-sick/economically inactive target group (Annual Population Survey 2019)

| Study | Page | Population health condition/s | Population inactivity group | Group | Educational attainment | Previous employment history | Comparison to UK long-term-sick/economically inactive population (APS 2019, target group) | Judgment |
| --- | --- | --- | --- | --- | --- | --- | --- | --- |
| 1.Craig et al., 2014 (ENDEAVOR) | p. 147 | Early psychosis | Long-term worklessness. | Intervention (IPS+MI): Midlands 1 (n=42) | Not reported | Midlands 1 (n=42): Ever worked 83%<br>London 1 (n=39): Ever worked 67%:<br>weighted average: 75% | Ever-worked rate (75%) is 2pp below the 77% APS 2019 comparator (never worked 23%). | Similar (employment); no education data |
| 2.Heslin et al., 2011 (SWAN) | p. 135 | Severe mental illness | Long-term worklessness. | Intervention (IPS, n=109) | Not reported | Worked in last 5 years: 51% | Worked-in-last-5-years rate (51%) is 29pp above the 22% APS 2019 'worked within last 5 years' comparator (worked within 2 years 9% + worked 2-5 years ago 14%) - markedly less disadvantaged on recent employment history. | Less disadvantaged (employment); no education data |
| 3.Moran, 2017 (2015 ESA Reform Trials Synthesis) | pp.16-17, 21, 56, 80-81, 83 | Mental health and physical health, , substance use disorders | Long-term benefit dependency (Incapacity Benefit/ESA), multiple health-related barriers, and long-term economic inactivity. | Voluntary Early Intervention (VEI, n=7,865) | Not reported | Not reported | Probably similar as IB/ESA client recipients. | Similar based on client group description. |
| 4.Howard et al., 2010 (SWAN, 1-year follow-up) | p. 406 | Severe mental illness | Long-term worklessness. | Intervention (IPS, n=109) | Not reported | Worked in the past 5 years: 51% (56/109) | Worked-in-last-5-years rate (51%) is 29pp above the 22% APS 2019 'worked within last 5 years' comparator - markedly less disadvantaged on recent employment history. Note: this is the same SWAN trial cohort as Heslin et al., 2011 (2-year follow-up of the same study) - the baseline table is identical, not independent data. | Less disadvantaged (employment); no education data |
| 5. Burns et al., 2015 (IPS-LITE trial) | p. 353 | Severe mental illness | Long-term worklessness. | IPS-LITE (n=62) / Standard IPS (n=61) | IPS-LITE: General education: median 13 yrs [IQR 12.0-14.0]; entered tertiary education 47%<br>Standard IPS: General education: median 13 yrs [IQR 11.0-13.0]; entered tertiary education 33% | Worked previously: 97% | IPS-LITE: Tertiary-entry rate (47%) is 21pp above the 26% APS 2019 A-level-or-higher comparator. Ever-worked rate (97%) is 20pp above the 77% APS 2019 comparator.<br>Standard IPS: Tertiary-entry rate (33%) is 7pp above the 26% APS 2019 comparator (within the similar band). Ever-worked rate (97%) is 20pp above the 77% APS 2019 comparator. | Less disadvantaged (education and employment) |
| 6. Lindley et al., 2015 (Want2Work) | p. 644 | Mixed (Incapacity Benefit recipients) | Long-term welfare benefit receipt, worklessness, and residence in economically weak, depressed local labour markets in Wales. | Want2Work participants (treated, unmatched, n=6424) | No qualifications 50%; NQF1 15%; NQF2 20%; NQF3 9%; NQF4 2%; NQF5 4% | Never worked 5%; worked within last 2 years (0-24 months) ~43% | No-qualifications rate (50%) is 13pp above the 37% APS 2019 comparator - more disadvantaged educationally. Never-worked rate (5%) is 18pp below the 23% APS 2019 comparator, and the worked-within-2-years rate (~43%) is 35pp above the APS 2019 comparator (9%) - markedly less disadvantaged on both lifetime and recent employment history. | Mixed: more disadvantaged (education), less disadvantaged (employment) |
| 7. Major et al., 2010 | p. 4 | First episode psychosis | Mixed | Whole cohort (n=114); not split by intervention vs comparison area | Basic secondary only 55%; A-levels 17%; non-degree professional qual. 7%; university (not completed) 13%; degree 6%; postgraduate 2% | Prior employment history: 75% previously employed; baseline: 67% unemployed, 14% competitively employed, 16% in education, 3% housewife/househusband. | Degree/postgraduate rate (8%) is 0pp above the 8% APS 2019 'degree or equivalent' comparator - essentially identical. Never-employed rate (25%) is 2pp above the 23% APS 2019 never-worked comparator - essentially matching, despite this being a young first-episode-psychosis cohort (mean age 24). | Similar (education and employment) |
| 8.Adams et al., 2022 (Employment Advisers in IAPT) | pp.42-48 (Tables 2.2-2.4); pp.110, 141-143 (Table D.1, D3) | Common mental health problems (some with long-term physical health conditions) | Mixed | Looking-for-work subgroup (31% of sample) | Looking-for-work subgroup: Level 5+ 40%; Level 3-4 23%; None or up to Level 2 37%<br>Off-work-sick subgroup: Level 5+ 51%; Level 3-4 26%; None or up to Level 2 23% | Never worked 6%; 51% unemployed <12 months, 33% unemployed 12+ months or never worked (16% unrecorded). | Looking-for-work subgroup: Level 5+ rate (40%) is 32pp above the 8% APS 2019 'degree or equivalent' comparator, and the 'None or up to Level 2' rate (37%) is 24pp below the comparable APS 2019 'GCSE or below' bucket (no qualification 37% + GCSE A*-C 24% = 61%) - markedly less disadvantaged educationally (note: Level 5 is a broader band than 'degree', so this likely overstates the gap somewhat).<br>Off-work-sick subgroup: Level 5+ rate (51%) is 43pp above the 8% APS 2019 comparator, and 'None or up to Level 2' (23%) is 38pp below the comparable APS 2019 'GCSE or below' bucket (61%) - markedly less disadvantaged educationally (same Level-5-vs-degree caveat as above).<br>Never-worked rate (6%) is 17pp below the 23% APS 2019 comparator - less disadvantaged on employment history too. | Less disadvantaged (education and employment) |
| 9.Dawson et al., 2019 (ESA WRAG 18-24 month re-referral pilots) | pp.38-40 (HMRC P45 charts); pp.51-56 (Annex A, Tables A.1-A.3) | Mental and physical health conditions (people in the ESA WRAG) | Long-term benefit dependency (ESA WRAG claimants with 18-to-24-month prognoses), | HCP / WP / JCP pilot arms (pooled description; no per-arm percentages reported) | Not quantified: described qualitatively as a lower-paid, less-well-educated population; 6%-9% screened as having a basic skills need across arms | Not quantified: HMRC P45 records show HCP participants relatively more likely to have been in employment 1-2 years prior to recruitment than WP or JCP participants (no percentages given). | No percentage figures were reported for qualifications or employment history, so no quantitative comparison to the APS 2019 comparator is possible. But probably similar as IB/ESA client recipients. | Similar based on client group description. |
| 10.Newton et al., 2023 (Health-Led) | pp.27-31 (Tables | Mental health and physical health | Mixed | SCR In-Work (SCR IW) - in-Work | Total sample (pooled across all 3 arms, not broken down | SCR In-Work (SCR IW) - :76% always in paid employment in last 2 | SCR In-Work sample – unsurprisingly less disadvantaged. SCR Out-of-Work (SCR OOW): Pooled A-level-or- | Less disadvantaged (education - |
| Employment Trials) | 1.2-1.11), pp.27-31 (Tables 1.2-1.11); pp.79-80 (Table 6.3) pp.27-31 (Tables 1.2-1.11); pp.79-80 (Table 6.3) |  |  | support arm / SCR Out-of-Work (SCR OOW) / West Midlands Combined Authority Out-of-Work (WMCA OOW) | individually): Degree/higher degree/NVQ4-5 17%; HE below degree 6%; A-levels/NVQ3 19%; GCSE A*-C 29%; GCSE D-G 12%; other 5%; no formal qualifications 12% | years, 14% more than half the time, 10% less than half the time, 0% not in paid employment in last 2 years.<br>SCR Out-of-Work (SCR OOW): 5% always in paid employment in last 2 years, 20% more than half the time, 25% less than half the time (50% some employment in last 2 years); 50% not in paid employment in last 2 years.<br>West Midlands Combined Authority Out-of-Work (WMCA OOW): 3% always in paid employment in last 2 years, 16% more than half the time, 22% less than half the time (41% some employment in last 2 years); 59% not in paid employment in last 2 years. | higher-equivalent rate (Degree/HE/NVQ4-5 + HE-below-degree + A-levels/NVQ3 = 42%) is 16pp above the 26% APS 2019 comparator; no-qualifications rate (12%) is 25pp below the 37% APS 2019 comparator - less disadvantaged educationally<br>SCR Out-of-Work (SCR OOW):: Some employment in last 2 years' (50%) is 41pp above the 9% APS 2019 'worked within last 2 years' comparator, and 'not in paid employment in last 2 years' (50%) is 18pp below the 68% APS 2019 comparator - less disadvantaged on recent employment history.<br>WMCA-OOW: 'Some employment in last 2 years' (41%) is 32pp above the 9% APS 2019 comparator, and 'not in paid employment in last 2 years' (59%) is 9pp below the 68% APS 2019 comparator - less disadvantaged on recent employment history | pooled figure; employment) |
| 11.Kantar Public et al., 2023 (Work and Health Programme evaluation) | pp.iv, 40-46, 108-109 pp.iv, 40-46, 108-109, 112-113 | Physical and mental health conditions, sensory impairments | Job seekers allowance and universal credit claimants | Mandatory referral group (Long-Term Unemployed 24+ months, 27% of sample) Voluntary referral group (Disability + Early Access, | Mandatory referral group: Level 3+ 29%; Level 2 27%; Level 1 12%; Below Level 1 22% Disability + Early Access: Level 3+ 31%; Level 2 28%; Level 1 12%; Below Level 1 22% | Mandatory referral group :13% never been in paid employment; of those who have worked, 13% last worked <1 year ago, 27% 1-3 years, 31% 3-10 years, 19% 10+ years ago Disability + Early Access: 12% never been in paid employment; of those who have worked, 26% last | Mandatory referral group: Level 3+ rate (29%) is 3pp above the 26% APS 2019 A-level-or-higher comparator - within the similar band. Never-worked rate (13%) is 10pp below the 23% APS 2019 comparator - less disadvantaged on lifetime employment history (though 77% have been out of work for 1+ years since last working, consistent with the 24+ month long-term-unemployed eligibility criterion). | Similar /higher (education); less disadvantaged (employment) |
|  |  |  |  | 73% of sample) |  | worked <1 year ago, 24% 1-3 years, 26% 3-10 years, 16% 10+ years ago | Disability + Early Access: Level 3+ rate (31%) is 5pp above the 26% APS 2019 comparator - within the similar band. Never-worked rate (12%) is 11pp below the 23% APS 2019 comparator - less disadvantaged on lifetime employment history. |  |
| 12.Bewley et al., 2009 (Pathways to Work, April 2006 expansion areas) | pp.17-18 (Table 2.2); p. 21 (Table 3.1); p. 55 (Table A.2) | Mixed | Incapacity benefit claimants. | Post-intervention cohort: incapacity benefits customers subject to Pathways (April 2006 areas) | Degree or equivalent 15%; A-level or equivalent 16%; GCSE or equivalent 29%; other qualification 5%; no qualification 35% | Baseline prior-employment history not reported. (Outcome, not baseline: 26% in paid work at the 19-month follow-up interview, of whom 83% worked 16+ hours/week and 55% worked 30+ hours/week - this is a post-intervention result, not a baseline work-history measure, so it is excluded from the target-group comparison below.) | No-qualifications rate (35%) is 2pp below the 37% APS 2019 comparator - essentially matching. A-level-or-higher rate (31%) is 5pp above the 26% APS 2019 comparator - also within the similar band. No baseline employment-history percentage was reported for comparison. | Similar (education); no baseline Employment data reported |
| 13.van Veggel et al., 2015 (IPS implementation in Sussex) | pp.289-291 (Table 2, Table 3) | Severe mental illness | Long-term worklessness. | IPS cohort (secondary mental health service clients) | Not quantified: no qualification breakdown reported | Not quantified: no employment-history percentages reported (only that vocational goals included competitive employment, voluntary work, education/training, or job retention) | No percentage figures were reported for qualifications or employment history, so no quantitative comparison to the APS 2019 comparator is possible. | No quantitative data for comparison |
| 14.Marwaha et al., 2014 | p. 137 | Severe mental illness | Long-term worklessness. | Model A: existing staff trained (n=39)<br>Model B: dedicated employment specialist (n=67 referred; 57 analysed) | Model A<br>Up to basic literacy 47%; up to GCSE 39%; A-level or higher 14%<br>Model B: Up to basic literacy 35%; up to | Model A: Ever employed: 64%<br>Model B: Ever employed: 81% | Model A: A-level-or-higher rate (14%) is 13pp below the 26% APS 2019 comparator. Ever-employed rate (64%) is 13pp below the 77% APS 2019 comparator.<br>Model B A-level-or-higher rate (29%) is 3pp above the 26% APS 2019 comparator. Ever-employed rate | More disadvantaged (education and employment) |
|  |  |  |  | after excluding 10 already in work) | GCSE 36%; A-level or higher 29% |  | (81%) is 4pp above the 77% APS 2019 comparator - within the similar band. |  |

### Comparator and judgment method

*Judgment compares each row’s figures to the long-term-sick/economically inactive APS 2019 comparator above, treating a gap of under ∼7 percentage points as ‘Similar’. MORE DISADVANTAGED = notably worse than the target group on that measure (red). SIMILAR = within ∼7pp of the target group (amber). LESS DISADVANTAGED = notably better than the target group (green). MIXED = more disadvantaged on one measure, less disadvantaged on the other (orange). NO DATA / NOT COMPARABLE = grey*.

*Caveats: these are national averages, not matched samples, and the underlying measures differ across studies (e.g. "entered tertiary education" is not the same as "A-level or higher"; "ever worked" and "worked in the last 2/5 years" are different concepts). Several studies here recruited young, first-episode-psychosis cohorts, where lower lifetime employment is partly attributable to age rather than disadvantage. Comparisons should be read as indicative*.

|  | All Studies | Non IPS studies | IPS studies | Effective studies |
| --- | --- | --- | --- | --- |
| Less disadvantaged | 6 (50%) | 1 (20%) | 5 (63%) | 4 (57%) |
| More disadvantaged | 1 (8%) | 0 (0%) | 1 (13%) | 0 (0%) |
| Similar | 4 (33%) | 3 (60%) | 2 (25%) | 2 (29%) |
| Mixed | 1 (8%) | 1 (20%) | 0 (0%) | 1 (14%) |
| No data | 1 | 0 | 1 | 1 |

Comparator: people who are long-term sick and economically inactive (i.e. out of the labour force specifically because of long-term sickness/disability) based on tabulation of Annual Population Survey (APS) 2019 data using the variables as outlined in the R script below:

| **Qualification** | **Frequency**<br>**(weighted)** | **Percentage** |
| --- | --- | --- |
| Degree or equivalent | 126349 | 7.9 |
| Higher education | 73763 | 4.6 |
| GCE A-level equivalent | 222735 | 13.9 |
| GCSE grades A*-C or equivalent | 385558 | 24.1 |
| Other qualifications | 202352 | 12.7 |
| No qualification | 587656 | 36.8 |

| **Work_history** | **Frequency** | **Percentage** |
| --- | --- | --- |
| Worked within last 2 years | 193806 | 8.7 |
| Worked 2-5 years ago | 303411 | 13.6 |
| Not worked in last 5+ years | 1225478 | 54.7 |
| Never worked | 516306 | 23.1 |

#### R script

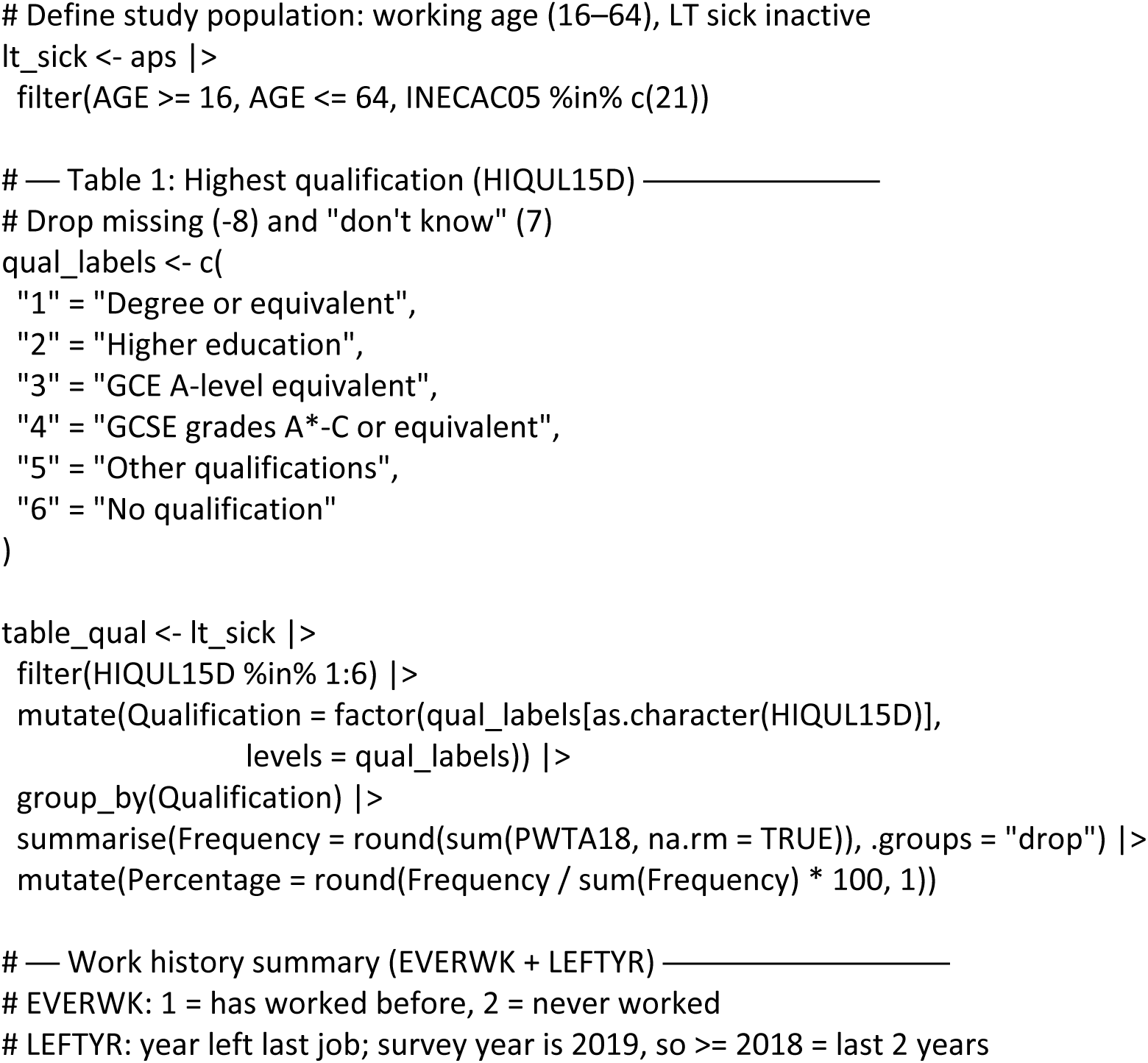
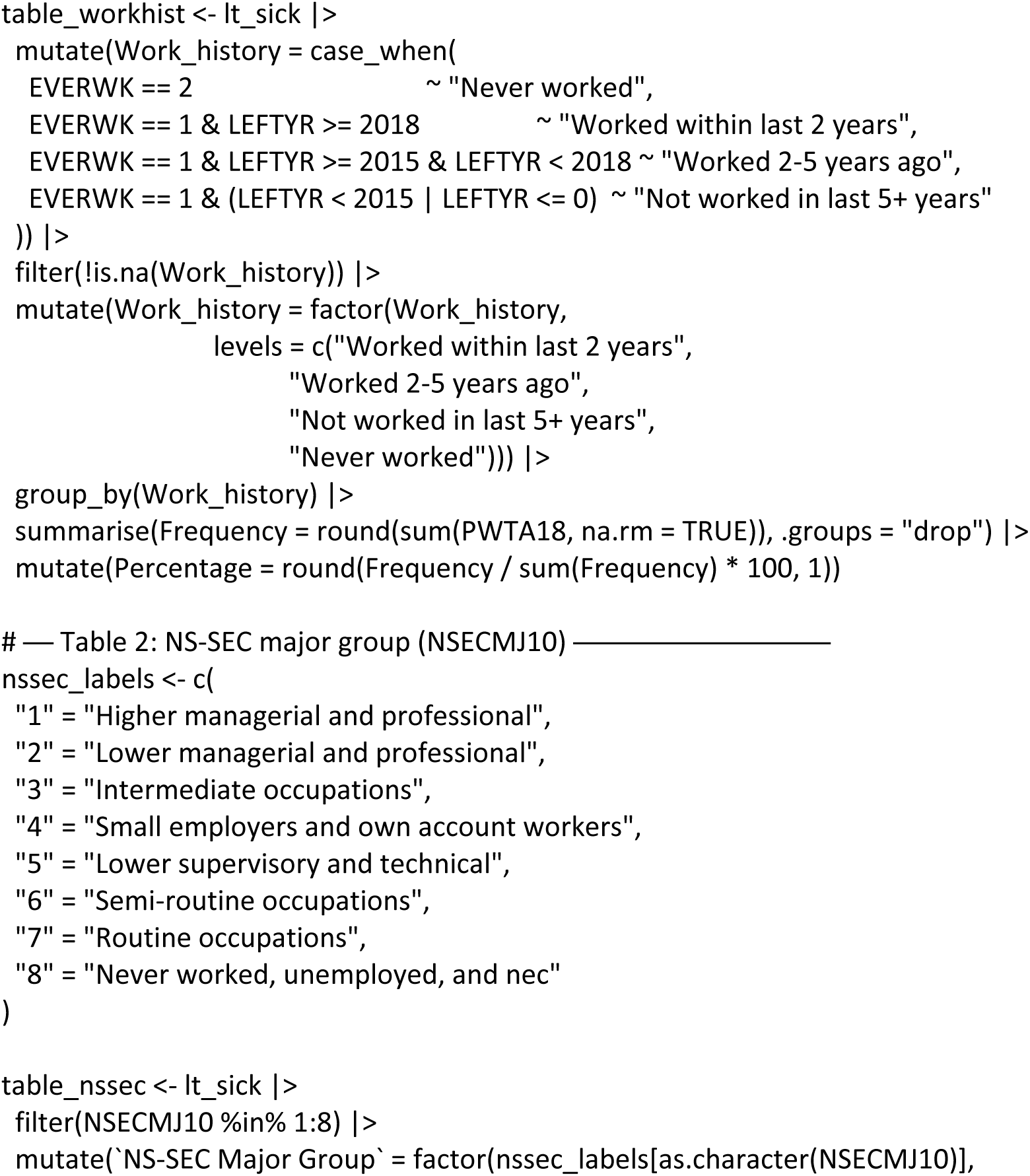
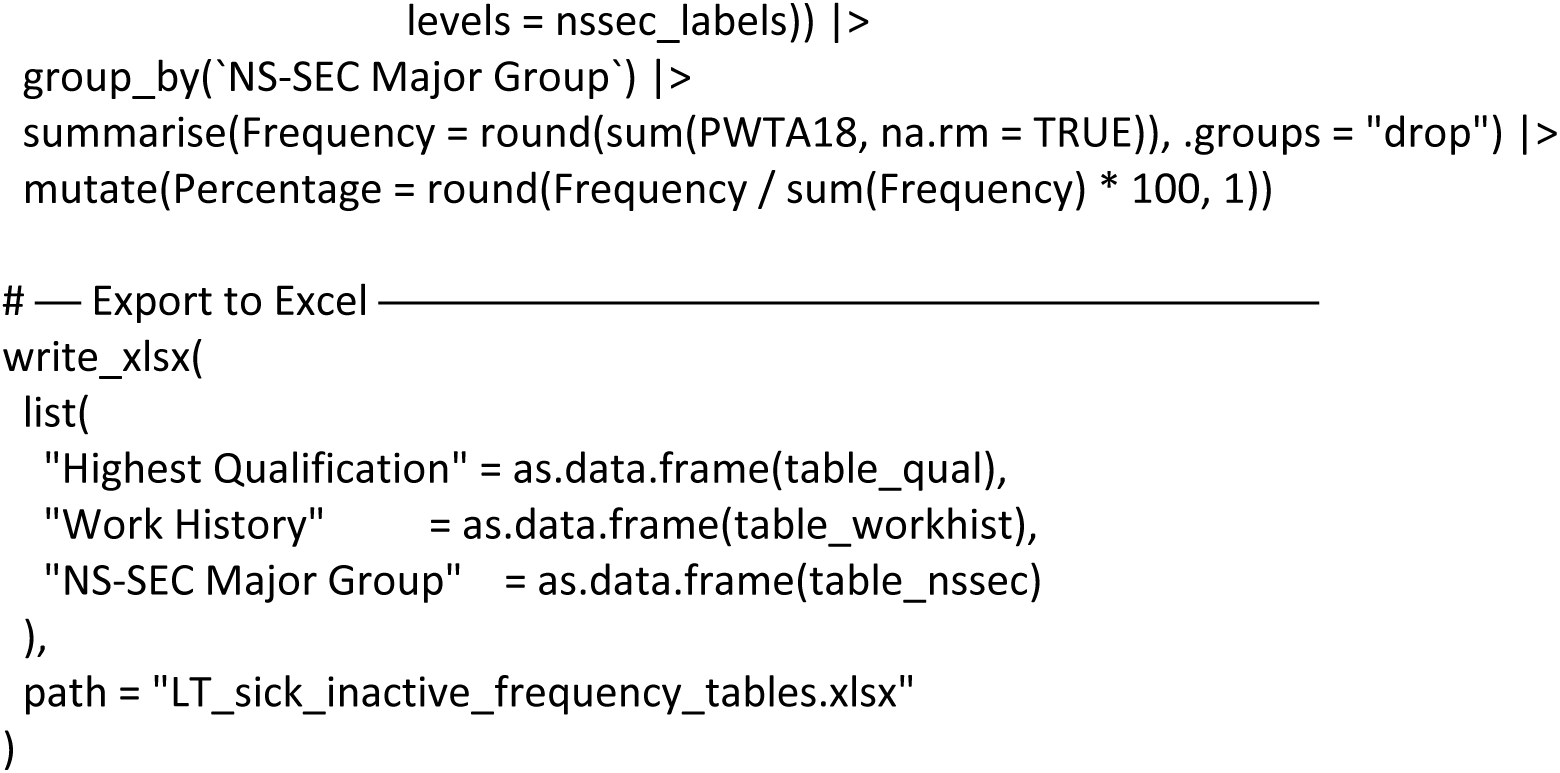

